# The pathophysiological mechanism of *Beni-koji Choleste-Help*/puberulic acid-induced kidney injury is proximal tubular mitochondrial dysfunction

**DOI:** 10.1101/2025.06.08.25329236

**Authors:** Yuta Sekiguchi, Makiko Mori, Haruka Maruyama, Yuki Nakao, Hiroaki Kikuchi, Shintaro Mandai, Fumiaki Ando, Koichiro Susa, Takayasu Mori, Yuma Waseda, Soichiro Yoshida, Yasuhisa Fujii, Wakana Shoda, Daiei Takahashi, Rei Okazaki, Kenji Ikeda, Tetsuya Yamada, Eisei Sohara, Shinichi Uchida, Yutaro Mori

**Author notes:** Lead contact and Corresponding author: Yutaro Mori, MD, PhD, FASN, Department of Nephrology, Graduate School of Medical and Dental Sciences, Institute of Science Tokyo, 1-5-45, Yushima, Bunkyo-ku, Tokyo 113-8510, Japan.

## Abstract

**Introduction:** In March 2024, kidney injury, sometimes with Fanconi syndrome, caused by a red yeast rice supplement (*Beni-koji Choleste-Help*), was reported in Japan. By November 24, 2024, 2,628 people had visited medical facilities, making it a social problem. Many patients still show decreased eGFR. However, the previous report only noted that puberulic acid, newly identified for its nephrotoxicity, was present in toxic lots. The pathophysiology of these nephropathies should be clarified. Here, we discovered that mitochondrial dysfunction in renal proximal tubular epithelial cells plays a pivotal role in the nephrotoxicity.

**Methods:** To assess the effects of *Choleste-Help* and puberulic acid, we performed RNA-seq, extracellular flux analysis (Seahorse XF Analyzer), immunofluorescence staining, Western blotting, and other assays across multiple models, including human kidney biopsy specimens, human-derived primary renal proximal tubular epithelial cells (hRPTECs), human renal organoids, and mice.

**Results:** A patient renal biopsy sample showed Kidney Injury Molecule-1 (KIM-1) expression in proximal tubules surrounded by activated myofibroblasts, indicating acute tubular damage and interstitial fibrosis. Mice treated with the toxic lot of *Choleste-Help* and puberulic acid showed kidney injury with Fanconi syndrome-like urinary findings. Pathological sections revealed tubular necrosis and interstitial fibrosis. RNA-seq analysis of whole kidneys showed that *Choleste-Help* and puberulic acid produced similar RNA patterns, suggesting puberulic acid is a causative agent. Gene Ontology (GO) analysis comparing the normal and toxic lot revealed downregulation of mitochondrial-related pathways. Puberulic acid also showed toxicity to hRPTECs and tubular organoids (tubuloids). *In vitro* experiments with hRPTECs revealed that it causes mitochondrial damage, especially to the mitochondrial respiratory chain, leading to cell death, mainly by necrosis.

**Conclusion:** Puberulic acid and a toxic lot of *Choleste-Help* cause direct mitochondrial damage to tubular epithelial cells, followed by necrosis.

**Graphical Abstract:** 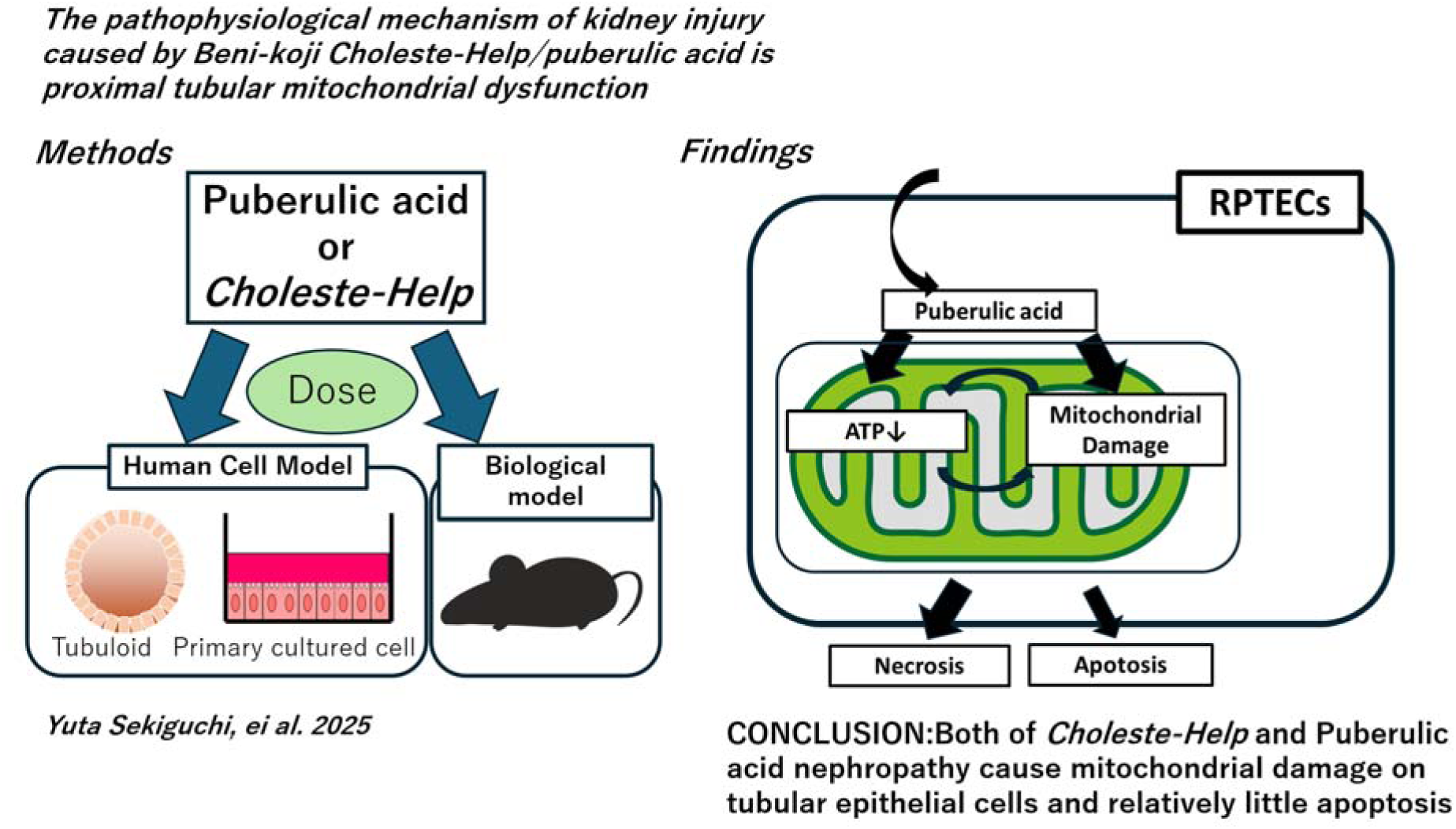

**Key Points:** First study shows puberulic acid causes mitochondrial injury preceding nephrotoxicity. First study shows matched renal transcriptomes for puberulic acid and Choleste-Help. Translational study using patient-derived primary cells and organoids for all assays.

## Introduction

In recent years, due to growing health consciousness, supplements and herbal medicines have been widely used. However, these often cause side effects.^1^ ^2^ A serious health issue arose in Japan from the second half of 2023 to 2024. Multiple cases of kidney dysfunction associated with Beni-koji tablets were reported, prompting Kobayashi Pharmaceutical Co., Ltd. to issue a voluntary recall on March 22, 2024.^3^ The supplement most taken by affected patients was *Beni-koji Choleste-Help* (*Choleste-Help*). According to Japan’s Ministry of Health, Labour and Welfare, 2,628 people consulted a doctor, and 348 were hospitalized due to renal injury after taking some specific Beni-koji supplements.^3^ The social significance of this study is extremely high. This supplement, which contains Monascus-derived red yeast^4^, was marketed as having lovastatin-like effects at lower doses.^5^ Patients who took these supplements developed kidney injury, proteinuria, and Fanconi syndrome,^6–8^ and many still had CKD-level kidney damage 120 days after starting treatment.^9^ A compound called puberulic acid and two other new compounds were identified as causative agents, with puberulic acid being the most likely.^10, 11^ However, only limited information is currently available.^12, 13^ Puberulic acid was originally developed as an antimalarial agent, but development was discontinued due to lethality observed in mice at high doses, the cause of which remains unknown.^14^ Aristolochic acid, the most widely known kidney-toxic substance from traditional Chinese medicine, is known not only to cause tubular damage but also to induce CKD and increase cancer risk.^15–17^ Therefore, it is essential to gather comprehensive information on this new toxic substance, including the affected pathways. In this report, we identify the main pathways of kidney damage caused by puberulic acid from a basic medical perspective. To the best of our knowledge, this is the first study to demonstrate puberulic acid toxicity in human cells.

## Methods

### Immunostaining of human renal biopsy samples

Informed consent was obtained from a patient with *Choleste-Help* nephropathy for publishing reports regarding renal biopsy and other clinical information. After deparaffinization with xylene, antigen retrieval was performed using Antigen Unmasking Solution (Citric Acid base, H-3300, Vector Laboratories Inc.). Details of the primary and secondary antibodies used in this study are listed in Supplementary Table.

### Animal treatment and tissue collection

Eight-week-old male C57BL/6J mice were randomly assigned to four groups (n = 6–10): (1) control (PBS), (2) *Choleste-Help* normal batch (1,000 mg/kgBW/day), (3) *Choleste-Help* toxic batch (1,000 mg/kgBW/day), and (4) puberulic acid (1 mg/kgBW/day) diluted with DMSO (0.1%). Treatments were administered via oral gavage or intraperitoneal injection for six days, followed by one day of rest. On day 7, mice were euthanized, and blood, urine, and kidneys were collected, snap-frozen in liquid nitrogen, and stored at −80°C. Hematological and urinary tests were performed by Oriental Yeast Co., Ltd.

### Histological Preparation and Staining

Hematoxylin and eosin (HE) staining was outsourced to the Research Core Center of the Institute of Science Tokyo. Masson’s trichrome staining was performed according to a standard protocol. One kidney section was selected per sample to minimize nonspecific blue staining. Whole-section images were acquired, and blue staining area was quantified using ImageJ Fiji. The percentage of blue-stained areas relative to the total kidney section was calculated for each sample.

### RNA-seq and its analysis

Mouse whole kidney total RNA was extracted using Sepasol (Nacalai Tesque, Japan) and purified with the Direct-zol RNA Miniprep Plus Kit (Zymo Research, R2070). Samples (n = 5, each) with RIN > 7.5 were processed by ReRexa Inc. using NEBNext kits and sequenced on NovaSeq X Plus. Reads were trimmed using fastp and quantified with Salmon (GENCODE vM33). DEGs were identified with DESeq2. PCA and heatmaps were generated in RStudio. Details of the methods are in the Supplementary Methods.

### Cell culture experiment

Human kidney samples were obtained from nephrectomy patients at the Institute of Science Tokyo Hospital, Tokyo, Japan. Primary human renal proximal tubular epithelial cells (hRPTECs) were isolated using a modified protocol.^18, 19^ Renal cortex tissue was minced and digested with Collagenase type II (1.0 mg/mL; Worthington Biochemical, NJ, USA). The reaction was stopped with fetal bovine serum, and cells were cultured in hRPTEC medium containing DMEM/F-12 and 1%BSA (Nacalai Tesque, Kyoto, Japan), Antibiotic–Antimycotic, hydrocortisone, and EGF (10ng/mL; Thermo Fisher Scientific, MA, USA), and ITS supplement (Sigma-Aldrich, MO, USA).

### Human renal tubuloid culture

hRPTECs were seeded onto ultralow attachment plates at a density of 5.0 × 10^5^ cells/well using Advanced RPMI 1640 medium (ThermoFisher Scientific) supplemented with 5% FBS. After a 2–3-day incubation period, Matrigel (Corning, NY, USA) was added to stimulate tubuloid formation. The following day, the culture medium was changed to Advanced RPMI 1640 medium with 5% FBS (Nacalai Tesque, Kyoto, Japan), EGF, FGF2, and HGF (10ng/mL of each) (tubuloids media). The media was changed three times a week to maintain optimal cell growth conditions. Tubuloids were usually ready for experimentation within 2 weeks of culture initiation.

### Western blot analysis (WB)

The samples were lysed, and the protein was purified using a lysis buffer [50 mM Tris-HCl (pH 7.5), 150 mM NaCl, 1% Nonidet P-40, 1 mM sodium orthovanadate, 50 mM sodium fluoride, and protease inhibitor cocktail]. The protein content of the Tuburoids was low; therefore, four wells under the same conditions were combined. Proteins were separated by 10% SDS-PAGE and transferred onto a nitrocellulose membrane. Protein bands were visualized using Western Blue (Promega, WI, USA). Densitometry was performed using ImageJ Fiji software.

### Cell viability assay

hRPTECs were seeded at a density of 2 × 10 cells/well in 96-well plates. After 2 days post-passaging, cells were treated with the indicated compound for 48 h. Cell viability was assessed using Cell Count Reagent SF (Nacalai Tesque, #07553) according to the manufacturer’s instructions. Absorbance was measured using a microplate reader (FilterMax F5, Molecular Devices). Cell viability was expressed as a percentage of absorbance relative to control. EC values were estimated using a four-parameter logistic model fitted to the viability data. Due to sparse data at higher concentrations, confidence intervals could not be fully resolved; therefore, the EC values should be interpreted with caution.

### Annexin V assay and flow cytometry

hRPTECs were seeded at a density of 5 × 10 cells//well in 12-well plates. After reaching confluence 2 days post-passaging, cells were treated with the indicated compound for 48 h. After treatment, cells were stained using the Annexin V Apoptosis Detection Kit (Nacalai Tesque, # 15342-54) according to the manufacturer’s instructions. Flow cytometric analysis was performed using a FACS Lyric™ system (BD Biosciences). Data were analyzed using FlowJo software (BD Biosciences), and cell populations were classified as live, early apoptotic, or late apoptotic/necrotic based on Annexin V and propidium iodide (PI) staining patterns.

### JC-1 staining

hRPTECs were seeded at a density of 5 × 10 cells/well in 12-well plates. After three days post-passaging, cells were treated with the indicated compound for 18 or 24 h. Mitochondrial membrane potential was assessed using JC-1 dye (T3168, Thermo Fisher Scientific) according to the manufacturer’s instructions. Briefly, cells were incubated with 1.5 μM JC-1 in culture medium at 37 °C for min. After incubation, cells were washed twice with PBS and analyzed by confocal microscopy (Eclipse Ti2, Nikon), with the NIS-Elements Advanced Research system. Fluorescence images were analyzed using ImageJ Fiji software. Three fields of view were randomly selected per well, and the entire field area was defined as the region of interest. The red-to-green fluorescence ratio was calculated to assess mitochondrial membrane potential.

### ATP measurement

hRPTECs were seeded at a density of 2 × 10 cells/well in 96-well plates. After reaching confluence 2 days post-passaging, cells were treated with the indicated compound for 6, 24, or 48 h. Cellular ATP levels were quantified using the CellTiter-Glo® 2.0 Cell Viability Assay (Promega, Madison, WI, USA) according to the manufacturer’s instructions. Luminescence was measured using a Varioskan™ LUX Multimode Microplate Reader (Thermo Fisher Scientific). Luminescence intensity, proportional to ATP levels, was recorded and normalized to control samples.

### Seahorse XF Cell Mito Stress Test

Mitochondrial respiratory function was assessed using the Seahorse XF Cell Mito Stress Test (Agilent Technologies) according to the manufacturer’s protocol. hRPTECs were seeded into Seahorse XFe24 Cell Culture Microplates at a density of 4 × 10 cells/well and cultured for 4 days. Cells were treated with puberulic acid (100 µM) for 4.5 or 9 h, with untreated cells as the control group. Culture medium was replaced with Seahorse XF assay medium (non-buffered DMEM supplemented with 10 mM glucose, 1 mM pyruvate, and 2 mM glutamine; pH 7.4) and incubated for 1 h at 37 °C in a non-CO incubator for equilibration. The assay was performed using a Seahorse XFe24 Analyzer. The following compounds were sequentially injected: oligomycin (1.5 µM), FCCP (4 µM), and rotenone (0.5 µM) plus antimycin A (0.5 µM), respectively, to determine non-mitochondrial respiration. Oxygen consumption rate (OCR) was measured at baseline and after each injection. Key parameters were calculated using Wave software (Agilent). All measurements were conducted in technical replicates (n = 6 wells per group), and cell-free wells were used for background correction.

### Quantification and statistical analyses

Figure legends indicate the number of samples assayed in each experiment. A one-way ANOVA followed by Tukey’s multiple comparisons test was used for general comparisons. When data were expressed as % of control, statistical comparisons were performed using Kruskal-Wallis test followed by Dunn’s post hoc test. A p-value < 0.05 was considered statistically significant. Asterisks indicate significance: * p < 0.05, ** p < 0.01, *** p < 0.001. All statistical analyses were performed using GraphPad Prism (GraphPad Software Inc., San Diego, CA, USA). To minimize the influence of aberrant values, extreme outliers with Z-scores > 2 were excluded as necessary.

### Study approval

Protocols for human kidney samples for primary culture and for analyzing biopsy samples were approved by the Institutional Review Board of the Ethics Committee of Institute of Science Tokyo (M2022-005, I2024-097) and conducted in accordance with the regulations of the ethics committee of the Kidney Disease and Transplant Center, Shonan Kamakura General Hospital. Animal experiments were approved by the Animal Care and Use Committee of Institute of Science Tokyo (A2023-109C3) in accordance with the animal experiment guidelines of the Japan Ministry of Education, Culture, Sports, Science, and Technology.

## Results

### A human kidney biopsy sample of a *patient with Cholesterol-Help showed KIM-1-positive* proximal tubular epithelial cells surrounded by α-SMA positive cells

Renal biopsy samples obtained from a patient who visited our affiliated hospital revealed marked tubular dilatation, vacuolar degeneration of tubular epithelial cells, and tubular casts in the lumen of the renal tubules (Figure 1a), whose case we reported recently.^7^ Immunostaining demonstrated the presence of KIM-1–positive tubular epithelial cells, an established marker of tubular injury,^20^ along with surrounding α-SMA expression, indicative of early fibrotic changes associated with tubular epithelial damage (Figure 1b). Although the degree of renal injury in this case was relatively mild, the findings suggest early-stage fibrosis.

**Figure 1.**
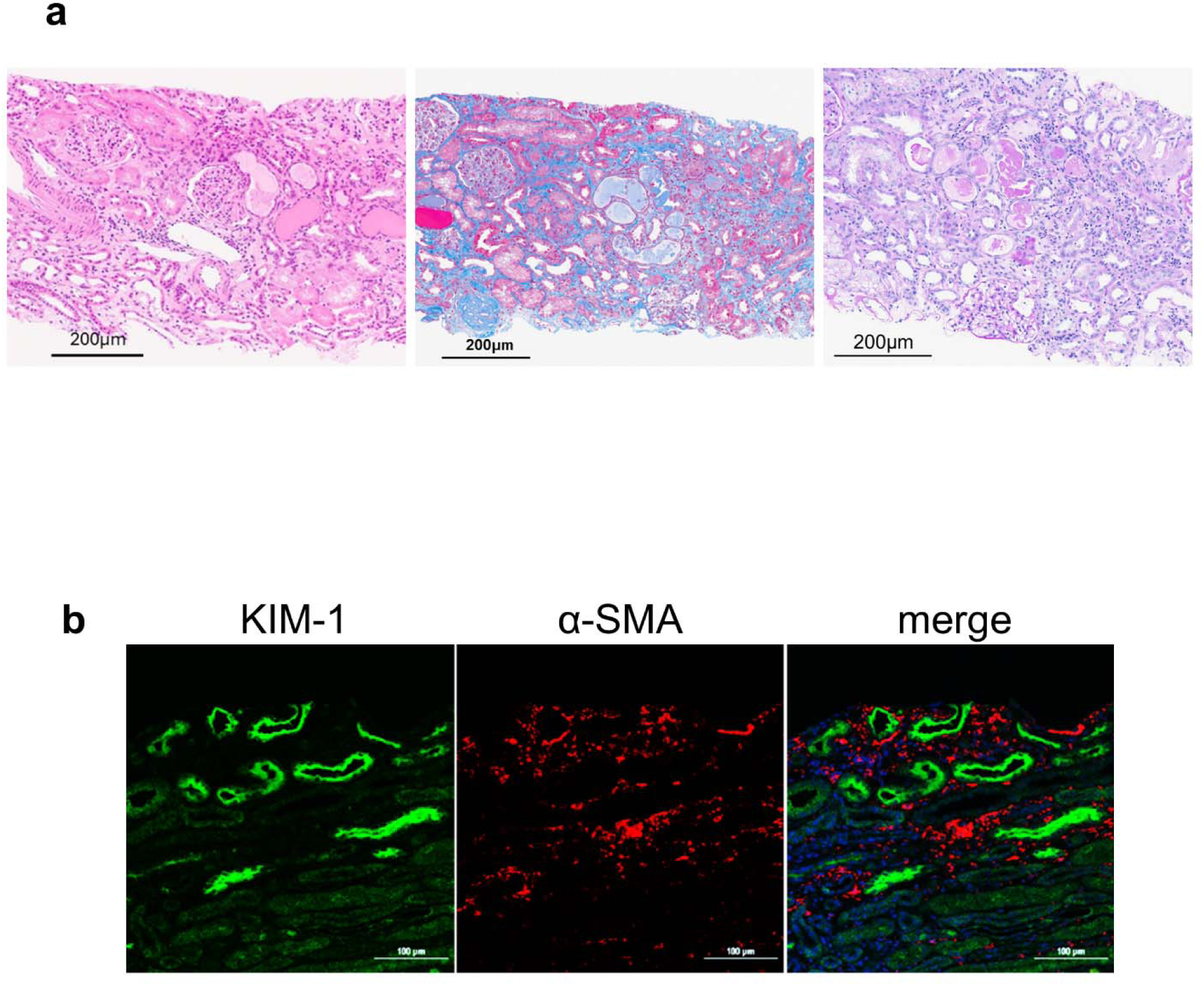
A human kidney biopsy sample from a patient with *Choleste-Help* Nephropathy shows renal tubular injury, necrosis, and surrounding interstitial fibrosis. (a) Hematoxylin and eosin staining, Masson’s Trichrome staining and Periodic Acid-Schiff staining of a human *Choleste-Help* nephropathy kidney. Scale bar: 200 μm. (b) Immunostaining of KIM-1 and α-SMA in a *Choleste-Help* nephropathy kidney. Scale bars: 100 μm.

### Acute kidney failure and tubular epithelial dysfunction in wild-type mice treated with *Choleste-Help* toxic lot and puberulic acid

To investigate *in vivo* toxicity and underlying mechanisms, we administered *Choleste-Help* (normal lot and toxic lot), the suspected causative compound puberulic acid, and vehicle (PBS) to mice once daily for six days. Following a one-day drug holiday, mice were sacrificed on day 7 (Figure 2a, Supplementary Figure 1). In both the *Choleste-Help* toxic lot and puberulic acid groups, serum biochemical analysis revealed elevated blood urea nitrogen (BUN) and creatinine (Cr) levels (Figure 2b). Urinalysis showed increased urinary albumin-to-creatinine (Alb/Cr), glucose-to-creatinine (Glu/Cr), and N-acetyl-β-D-glucosaminidase-to-creatinine (NAG/Cr) ratios, consistent with tubular injury and Fanconi syndrome. These findings indicate that both compounds induce proximal tubular injury, renal dysfunction, proteinuria in mice, like the features observed in human cases.

**Figure 2.**
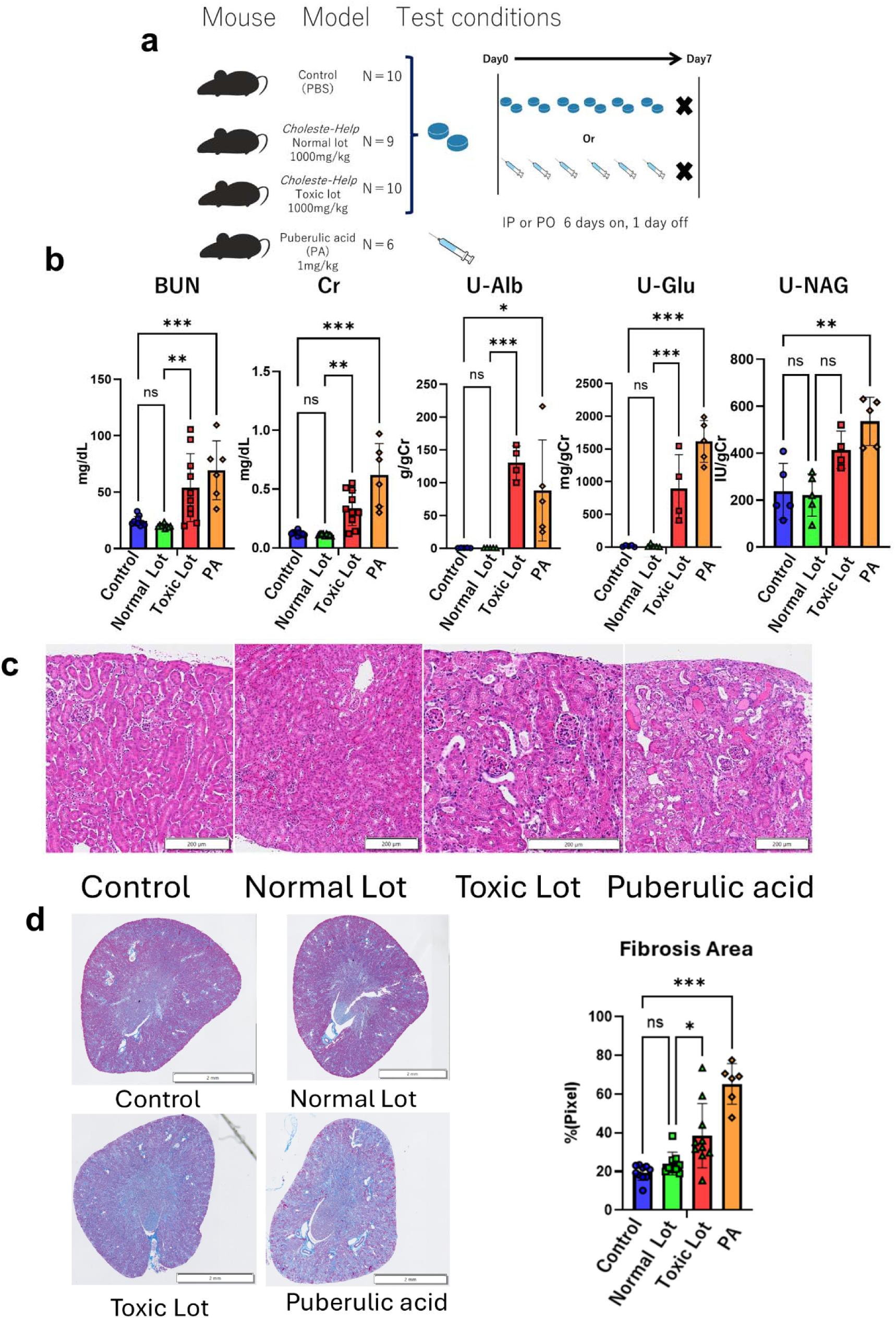
Acute kidney failure and tubular epithelial dysfunction consistent with Fanconi syndrome in wild-type mice treated with *Choleste-Help* toxic lot and puberulic acid. (a) Schematic overview of animal experiments treated with the *Choleste-Help* toxic lot or puberulic acid. (b) Blood urea nitrogen (BUN), serum creatinine (Cr), urinary albumin (U-Alb/U-Cr), urinary glucose (U-Glu/U-Cr), and urinary N-acetyl-β-D-glucosaminidase (U-NAG/U-Cr) in mice treated with PBS control, *Choleste-Help* normal lot, toxic lot, or puberulic acid. n = 10 for control and normal lot, n = 9 for toxic lot, and n = 6 for puberulic acid. P-values: BUN: 0.99 (Control vs Normal Lot), 0.0025 (Normal Lot vs Toxic Lot), 0.003 (Control vs Puberulic acid); Creatinine: 0.99, 0.004, <0.001; U-Alb/U-Cr: >0.99, <0.001, <0.001; U-Glu/U-Cr: >0.99, <0.001, h0.0137; U-NAG/U-Cr: 0.99, 0.0666, 0.0014. Bars represent mean ± SD. (c) Hematoxylin and eosin staining of mouse kidneys from PBS control, *Choleste-Help* normal lot, toxic lot, and puberulic acid. Scale bars: 200 μm. (d) Masson’s Trichrome staining and statistical analysis of fibrotic areas in mouse kidneys from PBS control (n=10), *Choleste-Help* normal lot (n=9), toxic lot (n =10), and puberulic acid(n=6). Scale bars: 200 μm. p-value for Control vs Normal Lot = 0.7659, Control vs Puberulic Acid < 0.001, Normal Lot vs Toxic Lot = 0.0288. Bars represent mean ± SD.

HE staining revealed tubular dilatation and vacuolar degeneration of tubular epithelial cells (Figure 2c), consistent with human biopsy findings. In addition, intratubular casts—also observed in human samples—were present in treated mice. Furthermore, Masson’s Trichrome staining demonstrated a significant increase in fibrotic areas in both the *Choleste-Help* toxic lot and puberulic acid groups (Figure 2d). These results suggest that the *Choleste-Help* toxic lot and puberulic acid can induce early-stage renal fibrosis *in vivo*.

### Mice treated with *the Choleste-Help* toxic lot and puberulic acid exhibited highly similar gene expression profiles, as revealed by RNA-seq analysis

We performed bulk RNA sequencing (RNA-seq) using whole-kidney samples (n = 5 for each sample) from the experimental groups as a transcriptomic analysis. Principal component analysis (PCA) and hierarchical clustering (Figure 3a, Supplementary Figure 2a) showed that the *Choleste-Help* toxic lot and puberulic acid groups clustered closely together, indicating highly similar transcriptomic responses.

**Figure 3.**
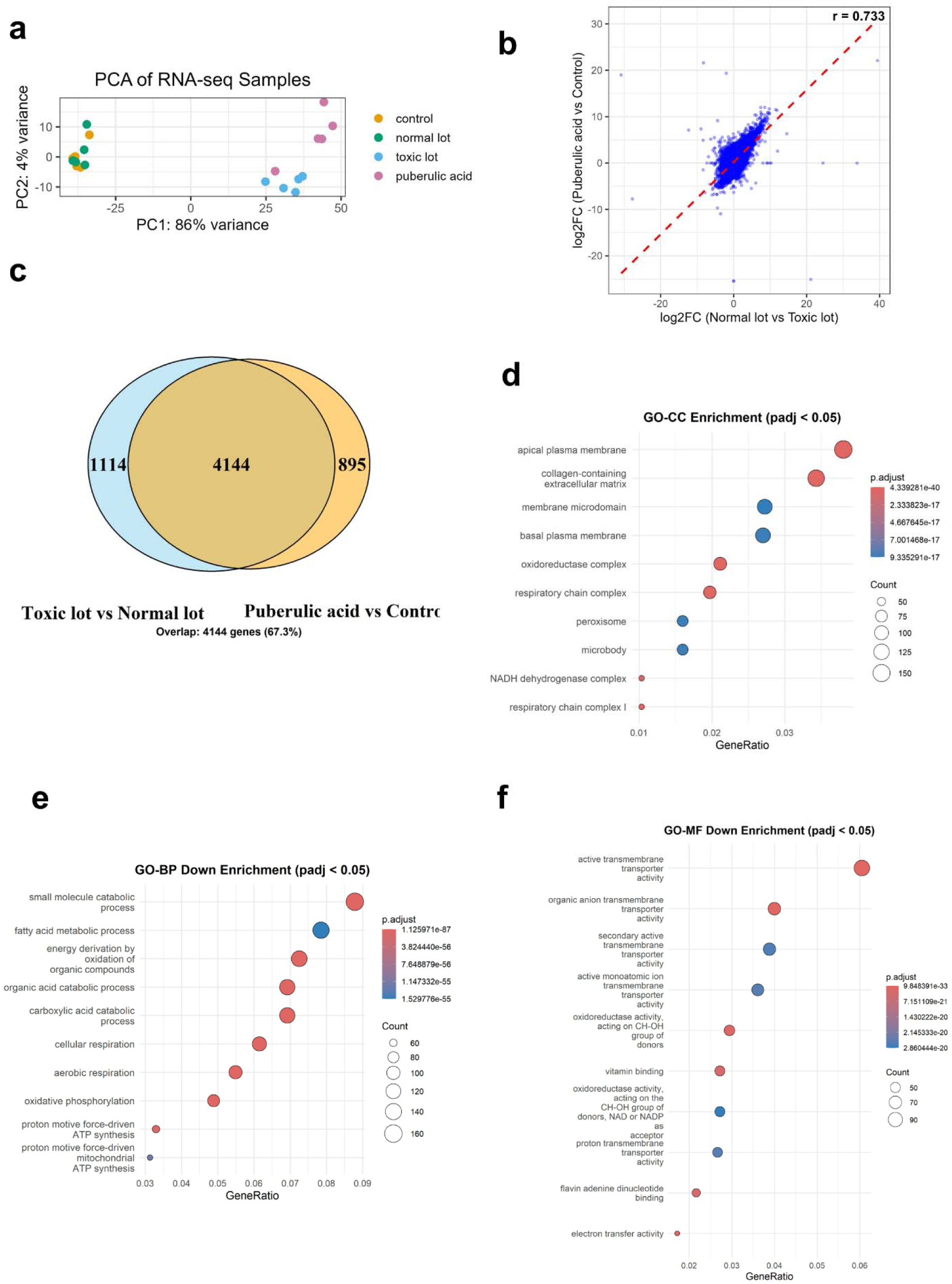
Mice treated with the *Choleste-Help* toxic lot and puberulic acid exhibit highly similar gene expression profiles, and mitochondrial disfunction occurs as revealed by RNA-seq transcriptomic analysis. (a) Principal component analysis (PCA) of RNA-seq samples using rlog-transformed gene expression values. Each point represents one biological replicate. Samples are color-coded by group: control (orange), normal lot (green), toxic lot (blue), and puberulic acid (magenta). PC1 and PC2 explain 86% and 4% of the variance, respectively. (b) Correlation of gene expression fold changes between comparisons. Each point represents a gene, with log2FC in normal vs toxic lot (x-axis) and puberulic acid vs control (y-axis). Pearson correlation coefficient: 0.733. The red dashed line indicates a linear regression fit. (c) Venn diagram showing the overlap of DEGs in puberulic acid vs control and toxic lot vs normal lot (padj < 0.05, |log2FC| > 1, baseMean > 10). A total of 4,144 genes were shared, representing 63.8% of the union set. (d) GO Cellular Component enrichment of downregulated genes. Dot plots show the top 10 enriched GO terms (padj < 0.05; log2FC < −1, baseMean > 10). The x-axis is the Gene Ratio. Color reflects adjusted p-values, and bubble size indicates gene count. (e) GO Biological Process enrichment of downregulated genes. Top 10 GO terms shown (padj < 0.05; log2FC < −1, baseMean > 10). Gene Ratio is shown on the x-axis. Color indicates significance; size shows gene count. (f) GO Molecular Function enrichment of downregulated genes. Top 10 MF terms shown (padj < 0.05; log2FC < −1, baseMean > 10). Dot size reflects gene count; color represents adjusted p-value.

To assess overall transcriptomic changes and data quality, a volcano plot and heatmap were generated to compare gene expression between the toxic and normal lots (Supplementary Figure 2b, c, d). These visualizations illustrate the global expression landscape and support the dataset’s reliability and robustness. In the volcano plot, *Spp1* and *Havcr1* were among the top upregulated genes, supporting the model’s validity in representing kidney injury.

Correlation analysis between differential gene expression profiles revealed a strong correlation (Pearson’s r = 0.736) between the comparisons of *Choleste-Help* toxic lot vs. normal lot and puberulic acid vs. PBS (Figure 3b). More than 65% of differentially expressed genes (DEGs) overlapped between the two comparisons, suggesting that the toxic responses induced by the *Choleste-Help* toxic lot and puberulic acid are highly similar at the transcriptomic level (Figure 3c). Although the precise concentration of puberulic acid in *Choleste-Help* is unknown and not equivalent to the dose used in the isolated puberulic acid group, the similarity in gene expression patterns strongly suggests that puberulic acid is likely a major contributor to the nephrotoxicity observed in the toxic lot of *Choleste-Help*.

### Mitochondrial disfunction occurred, as indicated by comprehensive transcriptomic analysis using RNA-seq

We performed Gene Ontology (GO) enrichment analysis comparing the *Choleste-Help* toxic lot and the normal lot. Based on GO cellular component analysis using the absolute value of log fold change, many of the top 10 downregulated GO terms were related to mitochondria, indicating they were among the most severely affected organelles (Figure 3d). GO terms associated with upregulated genes are shown in Supplementary Figure 3a. Notably, all mitochondrial-related terms showed a trend toward downregulation. Furthermore, in the GO biological process analysis, the top 10 downregulated pathways were predominantly associated with mitochondrial function (Figure 3e). GO terms for upregulated genes are shown in Supplementary Figure 3b.

In addition, GO analysis of the “molecular function” category revealed downregulation of various transporter activities, supporting the hypothesis that this compound induces a renal injury pattern consistent with Fanconi syndrome (Figure 3f). GO terms associated with upregulated genes are shown in Supplementary Figure 3c. Results of the GSEA and KEGG pathway analyses are presented in Supplementary Figures 4 and 5, respectively.

### Puberulic acid exhibited approximately half the cytotoxicity of cisplatin in hRPTECs and upregulated the DNA damage marker **γ**H2AX

To evaluate the potential nephrotoxicity of puberulic acid in humans, we exposed primary cultured hRPTECs to puberulic acid for 48 h and assessed cytotoxicity using a cell viability assay. Cisplatin was included as a positive control under the same conditions. Puberulic acid exhibited dose-dependent toxicity, with an EC of 66.04 μM—approximately twice that of cisplatin (EC = 31.24 μM)—indicating moderate cytotoxicity relative to cisplatin (Figure 4a).

**Figure 4.**
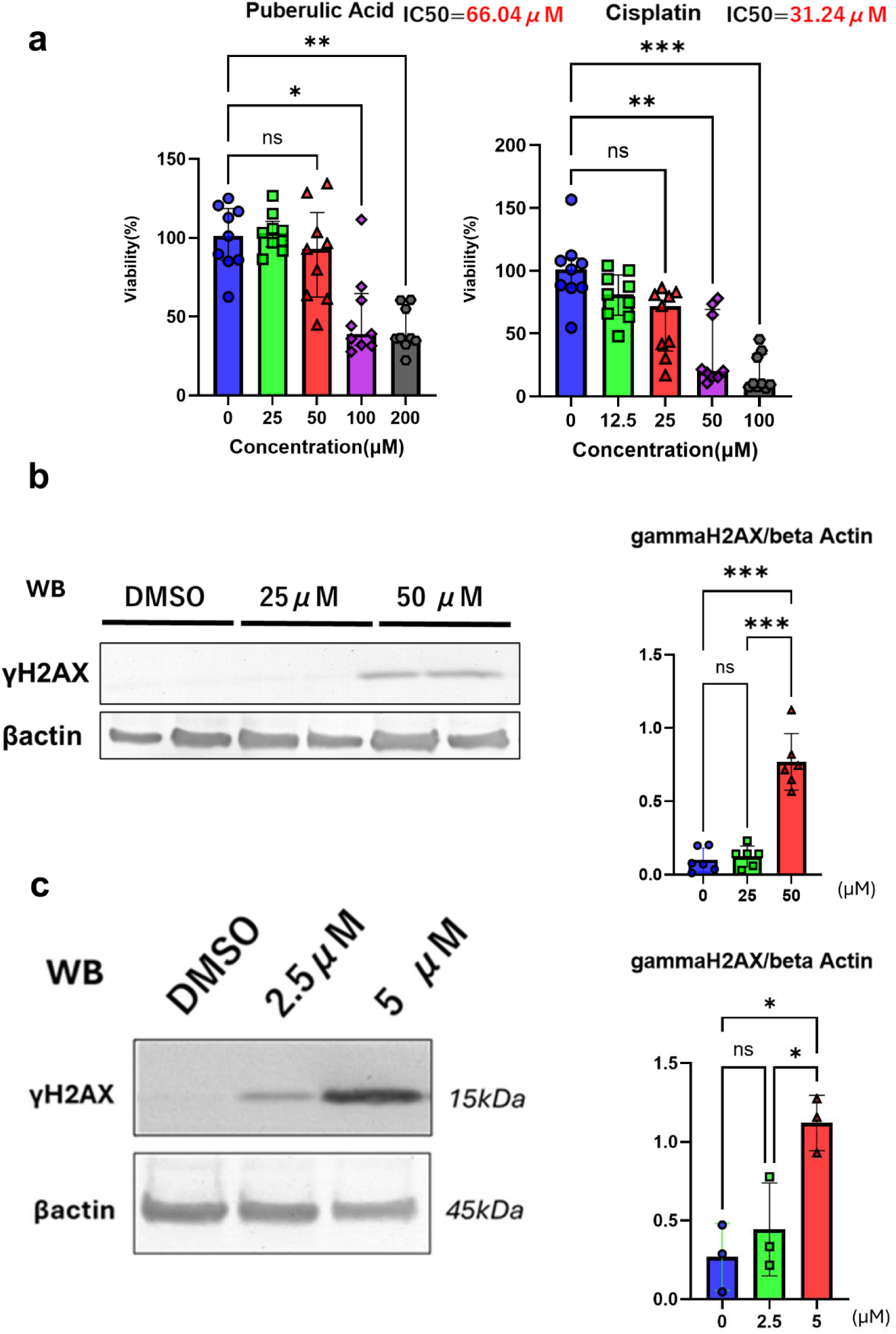
Puberulic acid exhibits about half the cytotoxicity of cisplatin in hRPTECs and upregulates kidney injury and DNA damage markers. (a) Cell viability of hRPTECs treated with puberulic acid or cisplatin (n = 9 per condition). Statistical test: Kruskal–Wallis. P-values vs control: >0.99, 0.0190, 0.0036 (Puberulic acid: 50 μM, 100 μM, 200 μM); 0.1699, 0.0025, <0.0001 (Cisplatin: 25 μM, 50 μM, 100 μM). Bars represent median ± IQR. EC estimated using a 4-parameter logistic model. (b) Western blot of γH2AX with beta Actin as a loading control from hRPTEC lysates. Densitometry values normalized to internal controls. n=6 from 3 independent experiments. P-values: 0.93 (0 μM vs 2.5 μM), <0.001 (2.5 μM vs 5 μM), <0.0001 (0 μM vs 5 μM). Bars represent means ± SD. Full blots in Supplementary Figure 10. (c) Western blots of γH2AX with beta Actin from puberulic acid-treated tubuloid lysates. Densitometry normalized to internal controls. n=3 from 3 independent experiments. γH2AX p-values: 0.65, 0.028, 0.010 (0 μM vs 2.5 μM, 2.5 μM vs 5 μM, 0 μM vs 5 μM). Bars represent mean ± SD. Full blots in Supplementary Figure 11.

The effects of puberulic acid were further examined using both conventional 2D cultures and 3D-cultured renal tubuloids derived from hRPTECs. Puberulic acid was administered at 48-h intervals for a total of three exposures. Western blot analysis revealed increased expression of the DNA damage marker γH2AX in both models following treatment (Figure 4b, c). Notably, the tubuloids responded to puberulic acid at approximately one-tenth the concentration required in the 2D cultures, indicating higher sensitivity in the 3D system.

DNA damage and its response are associated with cellular senescence through cell cycle arrest.^21^ The senescence-associated secretory phenotype is considered a driver of chronic fibrosis progression.^22, 23^ In mouse RNA-seq data, Cdkn1a expression was markedly upregulated, whereas Cdkn2a expression showed only a modest increase (Supplementary Figure 6a). Furthermore, in the *in vitro* model with a 6-day exposure, no induction of p16 expression was observed (Supplementary Figure 6b). These findings suggest that puberulic acid-induced cell cycle arrest occurs without clear evidence of cellular senescence.

### Annexin V assay revealed that necrosis accounted for majority of cell death

To evaluate the involvement of apoptosis, we performed live-cell immunostaining using Annexin V and propidium iodide (PI), followed by flow cytometric analysis. As a positive control, cells were treated with cisplatin, and the types of cell death were assessed at 48 h posttreatment.

The results showed that, under conditions with comparable numbers of viable cells, the proportion of apoptotic cells in the puberulic acid-treated group was approximately half that observed in the cisplatin-treated group, indicating a relatively low level of apoptosis (Figure 5a). Furthermore, at earlier points (24 and 40 h), the proportion of apoptotic cells in the puberulic acid group was even lower (Figure 5b). These findings suggest that puberulic acid induces apoptosis at relatively low levels. This observation is consistent with the renal biopsy findings in human cases, where necrosis was the predominant histological feature.

**Figure 5.**
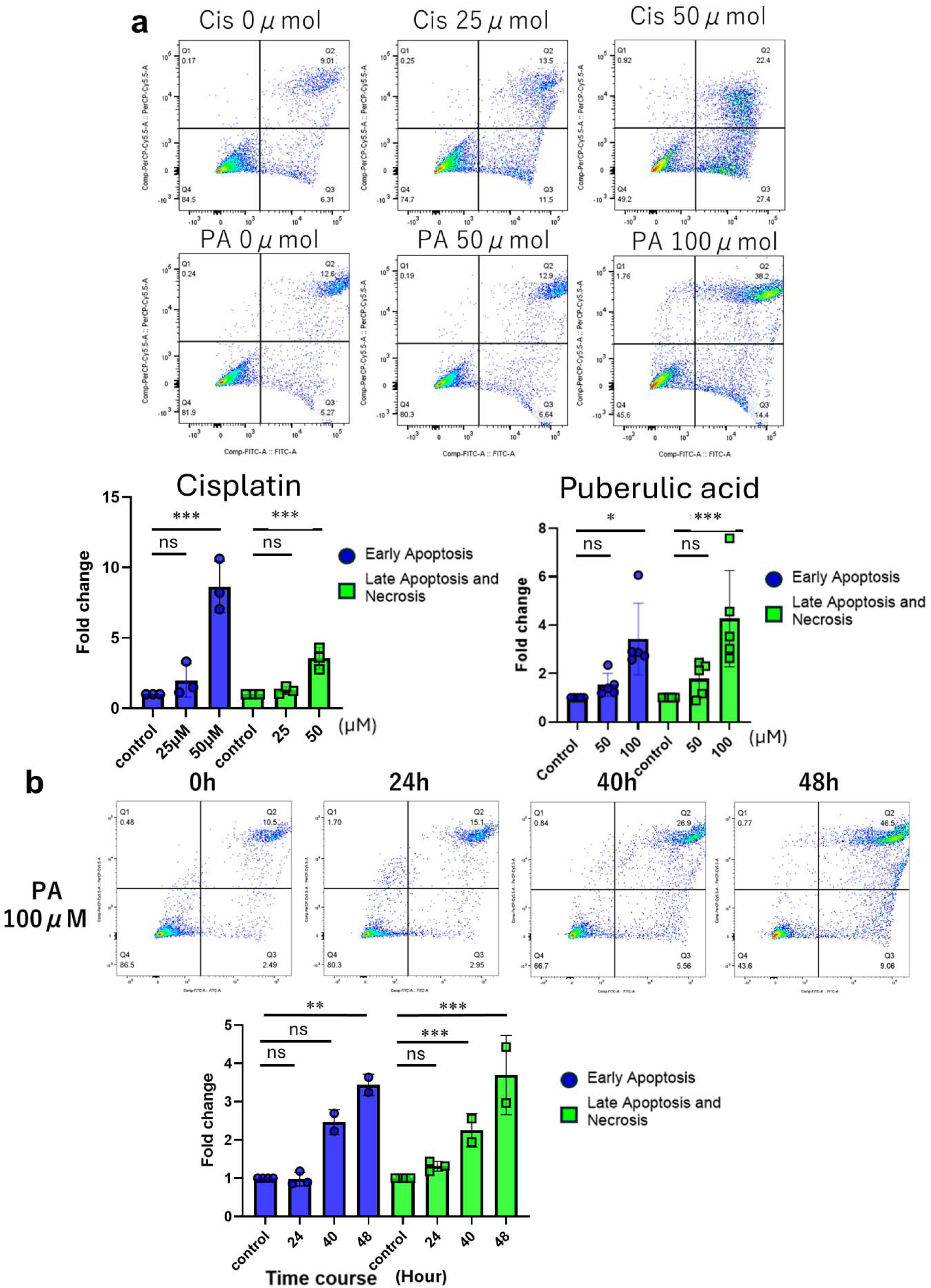
Necrosis accounts for majority of cell death in hRPTECs treated with puberulic acid. (a) Representative flow cytometry plots of hRPTECs treated with puberulic acid or cisplatin, stained with Annexin V and propidium iodide (PI) for the Annexin V assay. The graph shows fold change in early apoptosis (Annexin V positive, PI negative) and late apoptosis/necrosis (Annexin V positive, PI positive), normalized to the control group. Independent experiments were conducted three times for the cisplatin group (n = 3) and five times for the puberulic acid group (n = 5). Statistical analyses were performed on raw (non-normalized) data, and the graphs reflect these results. In the Cisplatin group, p-values for Early Apoptosis were 0.95 (0 μM vs 25 μM) and 0.0001 (0 μM vs 50 μM), and for late apoptosis/necrosis were 0.86 (0 μM vs 25 μM) and <0.001 (0 μM vs 50 μM). In the Puberulic Acid group, p-values for Early Apoptosis were 0.98 (0 μM vs 50 μM) and 0.020 (0 μM vs 100 μM), and for late apoptosis/necrosis were 0.16 (0 μM vs 50 μM) and <0.001 (0 μM vs 100 μM). Bars represent mean ± SD. (b) Representative flow cytometry plots of hRPTECs treated with 100 μM puberulic acid showing the time course of cell death. The graph shows fold change in early apoptosis (Annexin V positive, PI negative) and late apoptosis/necrosis (Annexin V positive, PI positive), normalized to the control group. Independent experiments were performed twice for both the 24-h group (n = 3) and the 40-h group (n = 2). Statistical analyses were performed on raw (non-normalized) data, and the graphs reflect these results. P-values for Early Apoptosis were 0.98 (0 μM vs 50 μM) and 0.020 (0 μM vs 100 μM), and for late apoptosis/necrosis were 0.16 (0 μM vs 50 μM) and <0.001 (0 μM vs 100 μM). Bars represent mean ± SD.

We also examined other forms of cell death, including necroptosis and pyroptosis (Supplementary Figure 7); however, we did not observe upregulation of pathway-specific molecules at the protein level in our *in vitro* experiments, suggesting that necroptosis and pyroptosis are not involved in puberulic acid-induced cell death.

In addition, at the 24-h time point, exposure to 100 μM puberulic acid induced only minimal cell death. Therefore, analyzing cells before this time point may provide further insights into the upstream mechanisms responsible for puberulic acid-induced cytotoxicity.

### JC-1 staining and ATP measurement revealed a decrease in mitochondrial membrane potential and ATP depletion before cell death onset

JC-1 staining was performed at 18 and 24 h following treatment with 100 μM puberulic acid—time points at which minimal cell death was observed (Figure 6a). At 18 h, a reduction in red fluorescence, indicative of healthy mitochondria, was already evident, accompanied by an increase in green fluorescence, reflecting a loss of mitochondrial membrane potential. This trend became more pronounced at 24 h.

**Figure 6.**
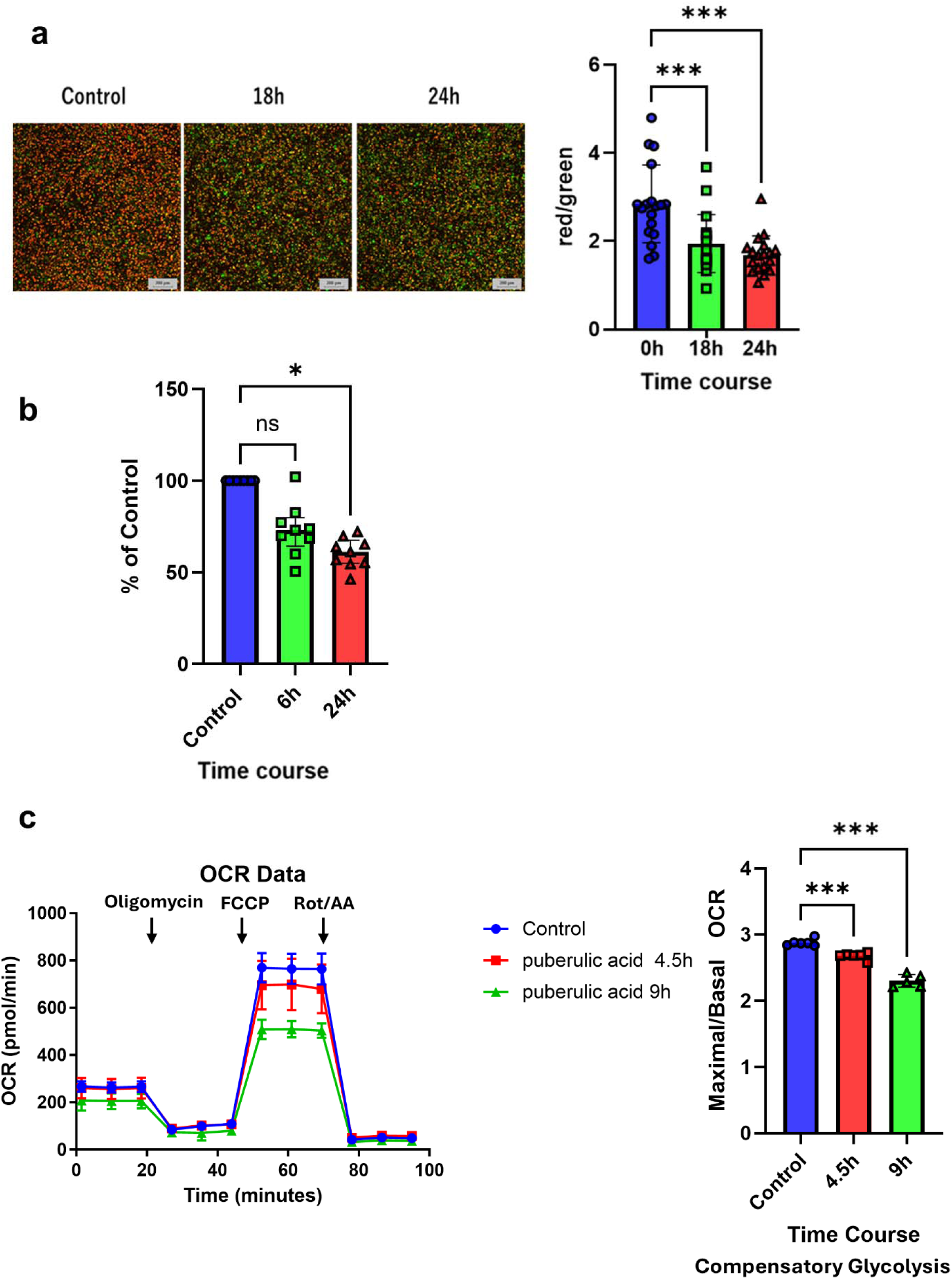
A decrease in mitochondrial membrane potential and ATP deficiency precede cell death, and mitochondrial respiration is suppressed following treatment with puberulic acid. (a) JC-1 staining of hRPTECs treated with puberulic acid and red/green ratio statistical analysis. Red and green indicate J-aggregates and JC-1 monomers, respectively. n = 18 in each condition. P-values were <0.001 (Control vs 18h), <0.001 (Control vs 24h). Bars represent mean ± SD. (b) ATP levels were measured and normalized to those of the respective control group at each time point. Values at 0 h represent theoretical references. n = 9 for each. Statistical test: Kruskal–Wallis. P-values were 0.45 (Control vs 6h), 0.012 (Control vs 24h). Bars represent median ± IQR. (c) The oxygen consumption rate (OCR) was measured in control cells and those treated with puberulic acid for 4.5 or 9 h using the Seahorse XF Cell Mito Stress Test. Each point represents the mean ± SD of six technical replicates. Statistical analysis was performed on the ratio of maximal to basal respiration (Maximal OCR/Basal OCR) to account for differences in cell number and to evaluate mitochondrial respiratory capacity in a normalized manner. One replicate in the 9-h group was excluded as an outlier (Z-score > 2). Puberulic acid time-dependently reduced maximal respiration, indicating progressive mitochondrial dysfunction. P-values were <0.001 (Control vs 4.5h), <0.001 (Control vs 9h).

These findings indicate that mitochondrial dysfunction occurs before the onset of cell death following puberulic acid exposure. Thus, puberulic acid-induced mitochondrial damage is likely one of the primary causes of its cytotoxic effects.

In addition, ATP measurement using a luminescence-based assay revealed a reduction as early as 6 h after treatment (Figure 6b). Puberulic acid impairs ATP production, likely through disruption of the mitochondrial respiratory chain.

### Seahorse assay revealed that puberulic acid causes reduction in maximal mitochondrial respiration

To evaluate the impact of puberulic acid (100 μM) on mitochondrial respiration, we conducted a Seahorse XF Cell Mito Stress Test and monitored the oxygen consumption rate (OCR) in control and treated cells (4.5 h and 9 h post-exposure). Puberulic acid exposure resulted in time-dependent suppression of mitochondrial respiration (Figure 6c). Maximal respiration (after FCCP injection) was significantly lower in the 9 h treatment group than in both the 4.5 h treatment and control groups, suggesting progressive impairment of the mitochondrial electron transport chain capacity. These results indicate that puberulic acid induces a progressive decline in mitochondrial respiratory function, potentially through cumulative damage to the oxidative phosphorylation machinery. A significant difference was also observed in ECAR at its peak following oligomycin injection (Supplementary Figure 10), indicating compensatory upregulation of glycolysis.

## Discussion

In this study, we demonstrated that *Choleste-Help* and puberulic acid induce similar patterns of renal injury at the transcriptomic level in mice. Importantly, we identified mitochondrial dysfunction—particularly in the mitochondrial respiratory chain—as a key mechanism contributing to puberulic acid-induced cell death in hRPTECs. We observed acute renal fibrosis in mouse models treated with puberulic acid or the toxic lot of *Choleste-Help*, as well as in human patients. This was supported by increased fibrotic areas on Masson’s trichrome staining and upregulation of fibrosis-related markers in RNA-seq analysis (Supplementary Figure 8, 9). We also observed proximal tubular epithelial cell death after puberulic acid treatment^7,9^ within a relatively short period, which was mainly necrosis. During proximal tubular epithelial necrosis, Damage-Associated Molecular Patterns are released from dead cells, resulting in myofibroblast activation and immune cell infiltration.^24, 25^ This may explain the early onset of fibrosis observed in our model.

Mitochondrial damage was demonstrated by early reductions in ATP levels and JC-1 signals in hRPTECs prior to cell death, supported by RNA-seq data from mice treated with puberulic acid or the toxic lot of *Choleste-Help*. Additionally, a marked reduction in mitochondria in electron micrographs of kidneys from puberulic acid-treated mice was reported in a preprint,^12^ consistent with our findings. As apoptosis is an ATP-dependent process,^26, 27^ necrosis tends to predominate under ATP-depleted conditions.^28, 29^ Therefore, we speculate that primary mitochondrial damage caused ATP deficiency, shifting the mode of cell death toward necrosis. Proximal tubular epithelial cells generate ATP mainly through mitochondrial oxidative phosphorylation, relying heavily on β-oxidation of fatty acids, with minimal glycolytic activity. In the proximal tubule, a large portion of solute reabsorption occurs via secondary active transport driven by Na /K -ATPase, which requires substantial ATP.^30^ Our Seahorse assay-based metabolic flux analysis revealed that puberulic acid disrupts mitochondrial respiration, supporting this ATP depletion mechanism. When mitochondrial dysfunction impairs energy production in proximal tubular cells, ATP depletion ensues, leading to Fanconi syndrome features, including glucosuria, aminoaciduria, phosphaturia, bicarbonate loss causing metabolic acidosis, and low molecular weight proteinuria.^31^ These are highly consistent with the Fanconi syndrome-like manifestations observed in both humans and mice after treatment with puberulic acid and *Choleste-Help*. We plan to investigate the detailed mechanisms of puberulic acid, including its uptake pathways and long-term effects.

In summary, we are the first to demonstrate that puberulic acid and *Choleste-Help* exhibit the same type of toxicity, which is mainly necrosis. We also showed that mitochondrial dysfunction precedes and likely contributes to this toxicity as a key mechanism. We hope that future studies building on this work will improve the prognosis and treatment of patients affected by such toxic effects.

## Supporting information

Supplementary things

## Data Availability

All data produced in the present study are available upon reasonable request to the authors

## Acknowledgment

We are deeply grateful to the generous study participants who donated resected kidney segments for this research.

## Author Contributions

YS performed the experiments, collected and analyzed the data, developed the experimental strategy, and prepared the manuscript. MM supported the experiments. HM, WS, and DT obtained human kidney biopsy samples from the patients. YS, YM, YN, and MM established the hRPTECs, and SM helped with the procedure. RO, KI and TY supported the seahorse assay. HK, FA, KS, TM, ES, and SU contributed to the data analysis. YW, SY, and YF resected the patients’ kidneys as standard treatment for malignant diseases. YM supervised the project, modified the experimental strategy, and finalized the manuscript. All authors discussed the results and implications and provided comments on the manuscript.

## Data Sharing Statement

The RNA-seq data generated in this study have been deposited in the NCBI Gene Expression Omnibus (GEO). The accession number is GSE301801. Other data supporting the findings of this study are available from the corresponding author upon reasonable request. Requests should be directed to Yutaro Mori, Lead Contact.

