## Supplementary things for "The pathophysiological mechanism of *Beni-koji Choleste-Help*/puberulic acid-induced kidney injury is proximal tubular mitochondrial dysfunction"

### Supplementary Methods

#### The detail of RNA-seq analysis

**RNA extraction, library preparation, and sequencing**: Total RNA was extracted by initially homogenizing kidney tissues in Sepasol-RNA I Super G (Nacalai Tesque, Japan), followed by purification using the Direct-zol RNA Miniprep Plus Kit (Zymo Research, R2070) according to the manufacturer's protocol. RNA integrity was assessed using an Agilent Bioanalyzer by ReRexa Inc, and all samples had RIN values greater than 7.5. RNA-seq library preparation and sequencing were also outsourced to ReRexa Inc. Libraries were constructed using the NEBNext Poly(A) mRNA Magnetic Isolation Module and NEBNext Ultra II Directional RNA Library Prep Kit. Sequencing was performed on an Illumina NovaSeq X Plus system, generating 2 × 150 bp paired-end reads (PE150), with approximately 6 giga bases of data and 40 million reads (20 million read pairs) per sample.

**Read preprocessing and quality control**: Raw reads were trimmed and filtered using fastp (v0.22.0) with the options: -q 20 -u 30 -n 10 -l 30 -w 8. Quality control was performed before and after trimming using FastQC (v0.12.1), and consolidated quality reports were generated using MultiQC (v1.28).

**Transcript quantification and annotation**: Transcript-level quantification was performed using Salmon (v1.10.3). The transcriptome index was built using GENCODE Mouse Release M33 (GRCm39), with the files gencode.vM33.transcripts.fa and gencode.vM33.annotation.gtf downloaded in April 2025 from https://ftp.ebi.ac.uk/pub/databases/gencode/Gencode_mouse/release_M33/. Gene-level count matrices were generated using the tximport package with the option countsFromAbundance = "lengthScaledTPM" to incorporate transcript length normalization, which improves compatibility with DESeq2's statistical model.

**Differential expression analysis**: Sample metadata (samples.csv) containing sample IDs and treatment groups was used as colData for differential expression analysis conducted using DESeq2 (v1.38.0). Genes with a total raw count ≤10 across all samples were excluded. Regularized log transformation (rlog) was applied for PCA. Differentially expressed genes (DEGs) were identified using an adjusted p-value (padj) < 0.05 and absolute log2 fold change > 1. For downstream GO enrichment analysis, genes were further filtered to include only those with a baseMean > 10. The primary comparison was made between the *Choleste-Help* toxic batch and normal batch. All downstream analyses including differential expression and GO enrichment were performed in RStudio running on Windows using the output files generated by Salmon in WSL2.

**Visualization**: Principal component analysis (PCA) was performed using the plot PCA function in DESeq2. The top 30 DEGs ranked by pad j were selected for heatmap visualization. Expression values were z-score normalized by genes, and heatmaps were generated using the pheatmap package (v1.0.12) with hierarchical clustering on both rows and columns.

**Reproducibility and data management**: Quantification steps up to Salmon were performed in a Linux environment using WSL2 (Ubuntu 22.04) with Miniconda (v24.9.2) for environment management. All downstream analyses including DESeq2 and enrichment analysis were conducted in RStudio on Windows.

### Antibody List

Primary Antibody

| Target Protein | Host Species | Supplier | Catalog Number | Dilution | Applicatio n |
|---|---|---|---|---|---|
| KIM-1 | goat | R&D Systems | AF1750 | 1:200 | IF |
| Alpha SMA | rabbit | Sigma-Aldrich | C6198 | 1:200 | IF |
| beta Actin | rabbit | Cell Signaling Technology | 4976S | 1:1000 | WB |
| phospho-Histone H2AX (Ser139) | rabbit | Cell Signaling Technology | 9718S | 1:1000 | WB |

Secondary Antibody

| Antibody | Host Species | Supplier | Catalog Number | Dilution | Applicatio n |
|---|---|---|---|---|---|
| anti-goat IgG | donkey | Thermo Fisher Scientific | A11055 | 1:200 | IF |
| antirabbit IgG alkaline phosphatase–conjugated | donkey | Promega | S373B | 1:3333 | WB |

**Supplementary Figures**

**Supplementary Figure 1.**

This figure presents the results shown in Figure 2band 2d, with the addition of a more appropriate control group (n=5) in which PBS was administered intraperitoneally to the mice, for comparison with the puberulic acid-treated group.

**Supplementary Figure 2.**

(a) Hierarchical clustering of RNA-seq samples based on Euclidean distance from rlog-transformed expression values. Clustering used complete linkage. The dendrogram illustrates transcriptomic similarities across control, normal lot, toxic lot, and puberulic acid. The x-axis labels indicate individual replicates. Shorter branches indicate higher similarity.

(b) Heatmap of the top 30 differentially expressed genes (DEGs) across treatments. Row-scaled Z-scores of normalized RNA-seq counts are shown (padj < 0.05, |log2 fold change| > 1, baseMean > 10). Each row is a gene, each column, a replicate. Samples are annotated by group. Genes and samples were clustered using Euclidean distance and complete linkage. Red and blue represent high and low expressions, respectively.

(c) Volcano plot of DEGs between toxic and normal lots. Each dot is a gene: red and blue indicate significant up- and downregulation (adjusted p < 0.05, |log2FC| > 1, baseMean > 10). Gray dots are non-significant. The top 10 up- and downregulated genes are labeled. Genes with |log2FC| > 10 (n = 6) were excluded. The total number of significant genes is shown.

(d) Volcano plot of differentially expressed genes between the puberulic acid group and control. Each dot represents a gene, with red and blue indicating significantly upregulated and downregulated genes, respectively (adjusted p-value < 0.05 and |log2 fold change| > 1, baseMean > 10). Gray points represent genes with no significant change. The top 10 upregulated and downregulated genes were labeled based on adjusted p-value ranking. For improved visualization, genes with extreme fold changes (|log2FC| > 10, n = 22) were excluded from the plot. The total number of significantly changed genes is indicated in the figure.

**Supplementary Figure 3.**

GO enrichment of upregulated genes. Dot plot showing the top 10 significantly enriched terms (padj < 0.05) among genes downregulated (log2FC < -1, baseMean > 10). The x-axis represents the Gene Ratio, defined as the proportion of input genes annotated to each GO term. Dot size indicates the gene count, while dot color represents the adjusted p-value.

**Supplementary Figure 4.**

GSEA of GO Biological Process. Gene Set Enrichment Analysis (GSEA) was performed using ranked log2 fold change values from differential expression analysis between toxic lot and normal lot. The dot plot displays significantly enriched GO Biological Process terms (padj < 0.05), with the gene ratio on the x-axis, and terms split by enrichment direction: "activated" (positive enrichment score) and "suppressed" (negative enrichment score). Dot size corresponds to the number of genes contributing to the term, and color reflects the adjusted p-value. Key suppressed pathways include mitochondrial and oxidative phosphorylation-related terms, whereas activated terms involve immune responses such as leukocyte activation and cytokine signaling.

**Supplementary Figure 5.**

KEGG pathway enrichment analysis of differentially expressed genes. Dot plot showing the top 10 significantly enriched KEGG pathways among upregulated (red, left) and downregulated (blue, right) genes in response to [Drug Name or Treatment]. Gene ratios (x-axis) indicate the proportion of DEGs involved in each pathway. The size of the dots represents the number of genes, and color intensity corresponds to the adjusted p-values. Pathways are ranked by statistical significance (padj) within each group.

**Supplementary Figure 6.**

(a) Expression levels of p16 (Cdkn2a) and p21 (Cdkn1a) across experimental conditions. Box plots show the log2-transformed normalized RNA-seq expression levels of Cdkn2a and Cdkn1a across four experimental groups: control, normal lot, toxic lot, and puberulic acid. Each box represents the interquartile range (IQR) with the median indicated by a horizontal line. Whiskers extend to 1.5×IQR, and outliers are omitted for clarity.
(b) This figure shows the Western blotting data of tubuloids treated with puberulic acid, focusing on the expression of p16, BAX, BCL-2, Cleaved Caspase3.

**Supplementary Figure 7.**

This figure shows the Western blotting data of hRPTECs treated with puberulic acid, focusing on the expression of cleaved Gasdermin D, cleaved Caspase-1, phospho-RIP3 and phospho-MLKL.

**Supplementary Figure 8.**

Expression of fibrosis-related genes in renal tissue. Boxplots showing the normalized expression levels ($log_2$ normalized counts + 1) of representative fibrosis markers (Acta2, Col1a1, Mmp2, and Tgfb1) across experimental conditions: control, normal lot, toxic lot, and puberulic acid. Notably, toxic lot and puberulic acid treatments resulted in a marked upregulation of Acta2, Col1a1, and Tgfb1 compared to control or normal lot, consistent with fibrotic responses.

**Supplementary Figure 9.**

KEGG pathway enrichment of fibrosis-related terms among upregulated genes following toxicant exposure.KEGG enrichment analysis was performed using genes with BaseMean > 10 that were significantly upregulated (adjusted $p < 0.05$) in the toxic lot group. Pathways relevant to fibrosis, including "ECM-receptor interaction", "TGF-beta signaling pathway", and "focal adhesion", were selectively enriched. The size of each dot represents the number of genes involved in the pathway, and the color indicates the adjusted *p*-value for enrichment. These results support activation of fibrotic signaling pathways at the transcriptomic level.

**Supplementary Figure 10.**

Extracellular acidification rates (ECARs) were measured in control and puberulic acid-treated cells using the Seahorse XF Cell Mito Stress Test at 4.5 or 9 h. Oligomycin, FCCP, and rotenone/antimycin A were sequentially injected at the indicated time points. Both puberulic acid groups exhibited elevated ECAR after oligomycin injection compared with the control group. Statistical analysis showed a significant ECAR increase at the post oligomycin peak, suggesting a compensatory glycolytic response to mitochondrial inhibition. Data are presented as mean ± SEM (n = 6 wells per group). P-values were <0.001 (Control vs 4.5h), <0.001 (Control vs 9h).

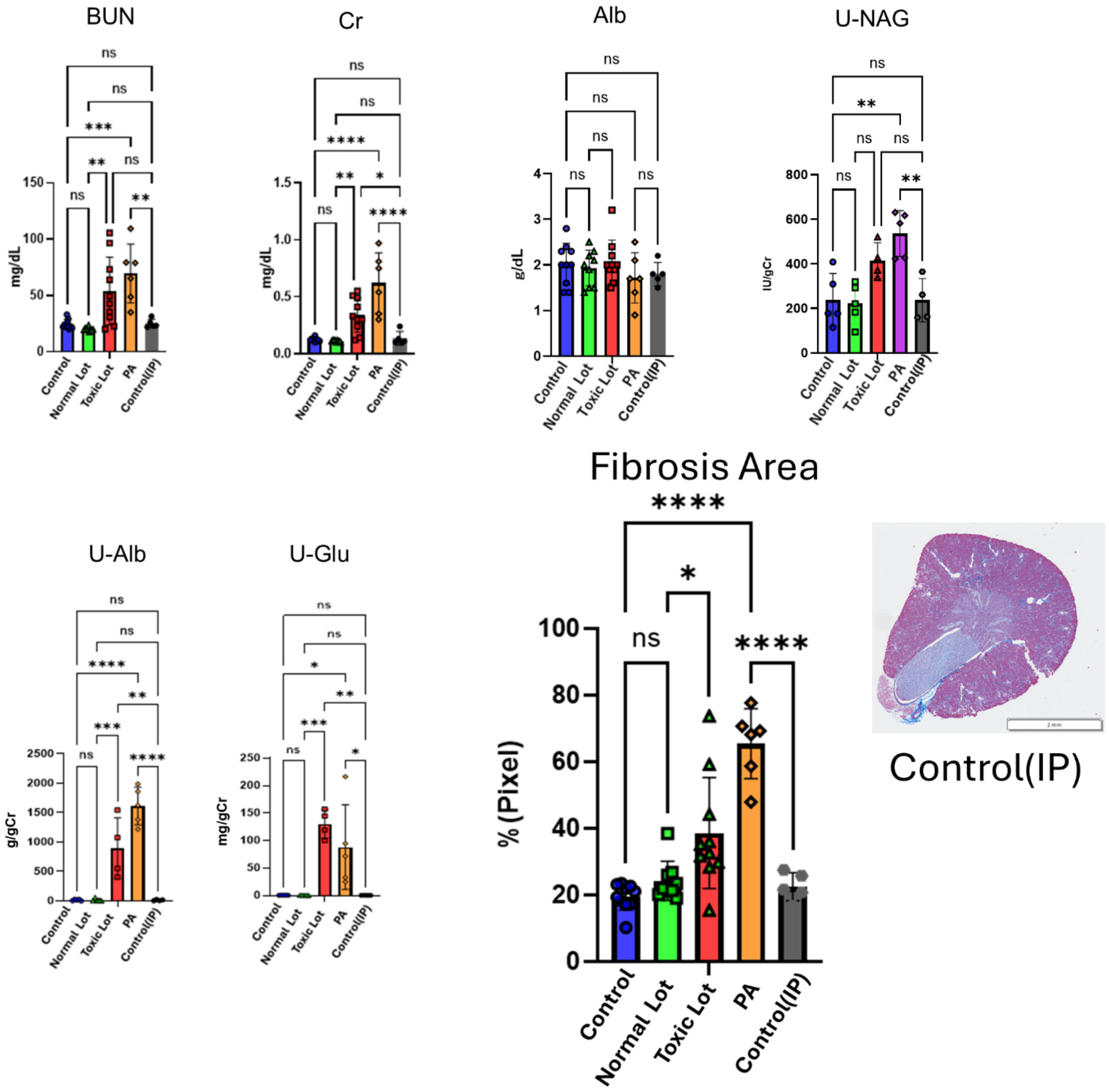


Supplementary Figure 1

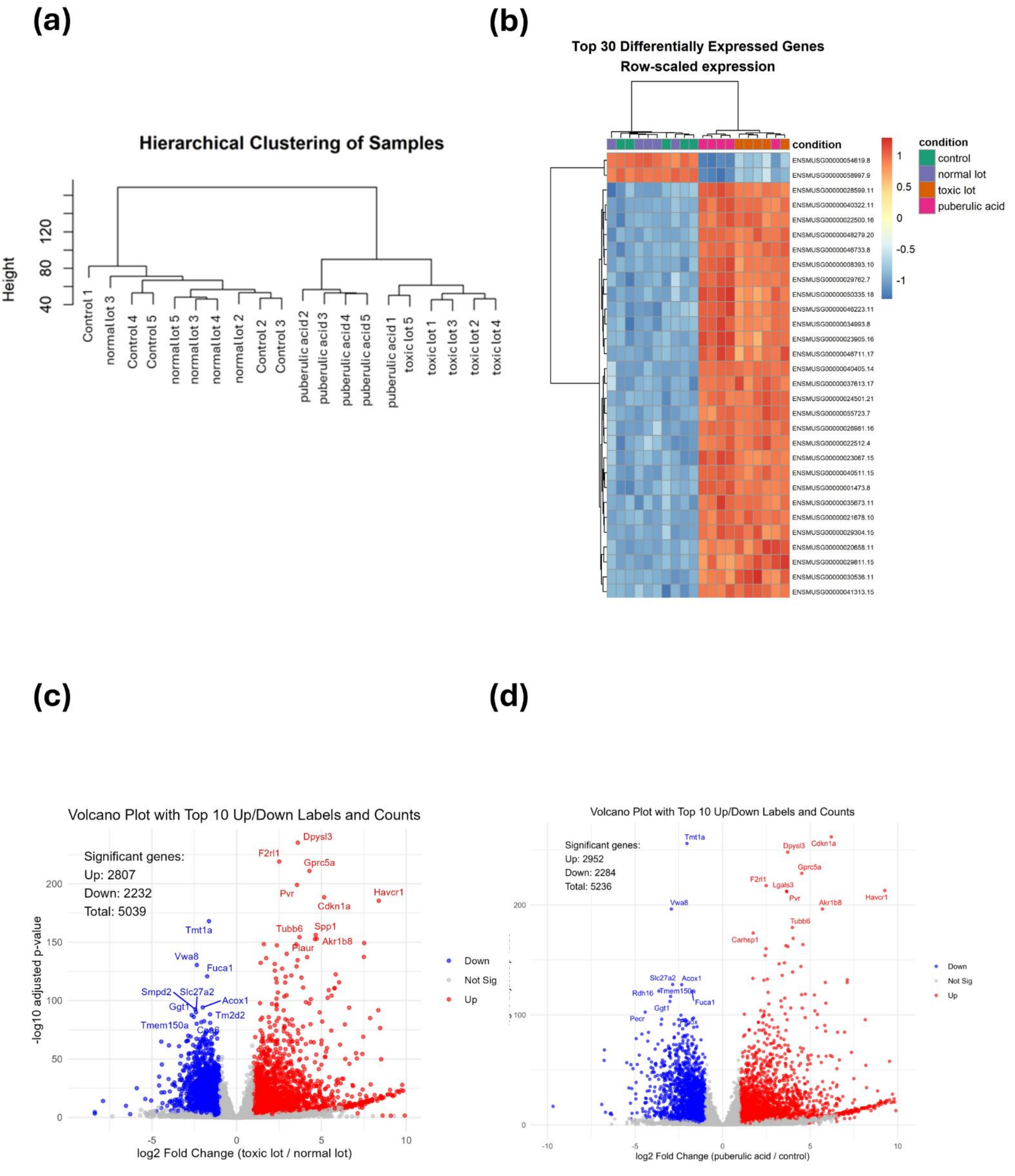


Supplementary Figure 2

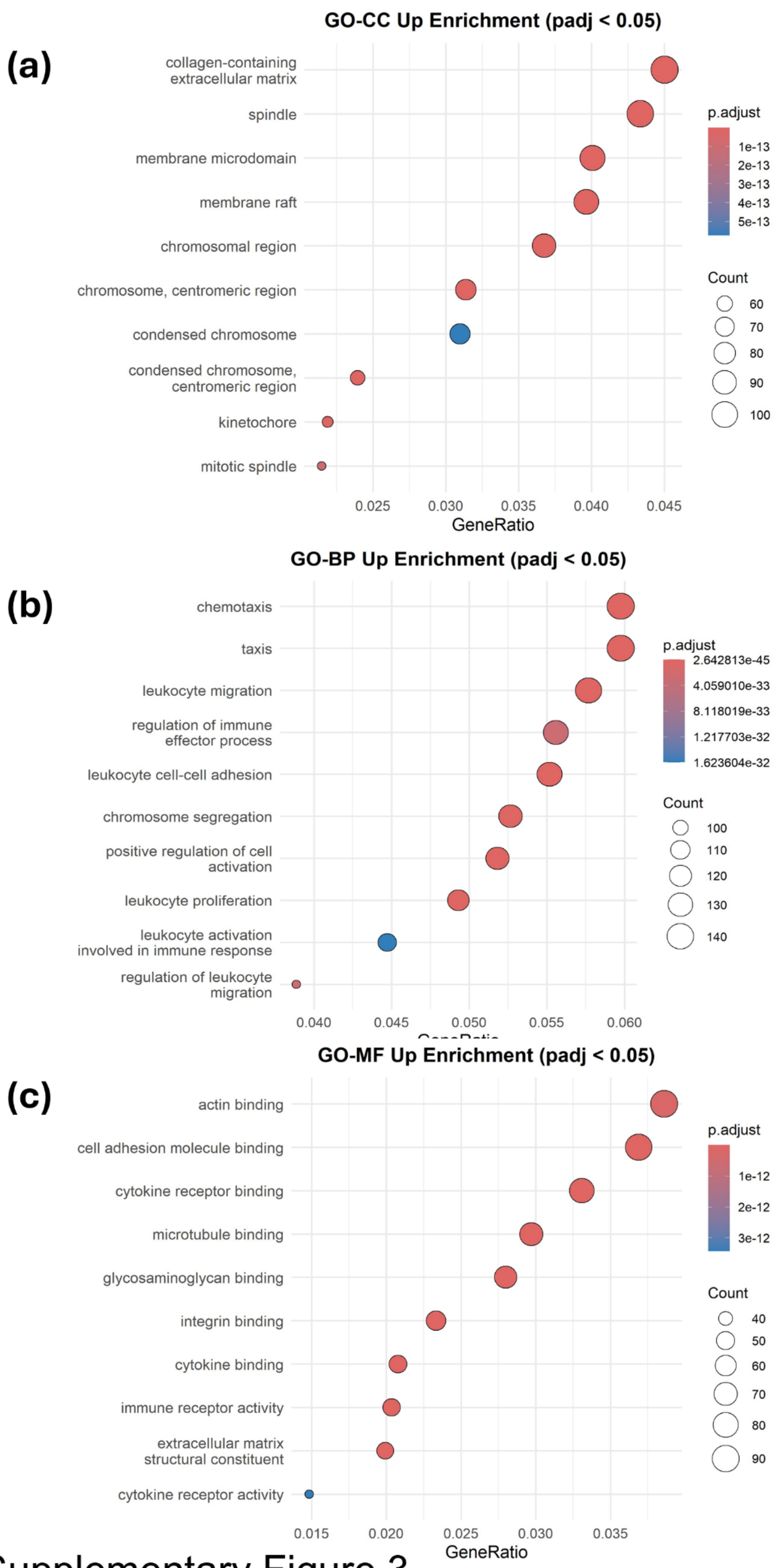


Supplementary Figure 3

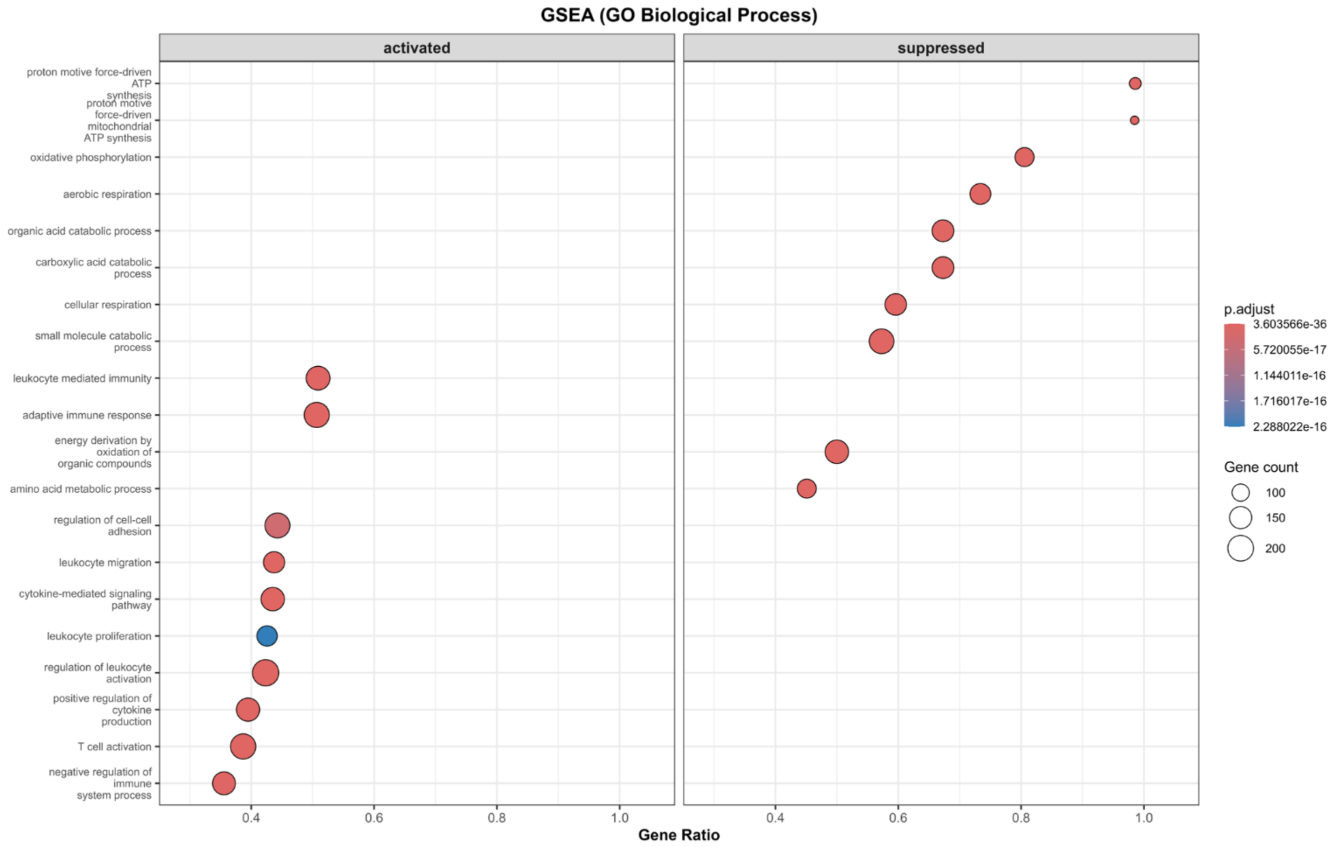


Supplementary Figure 4

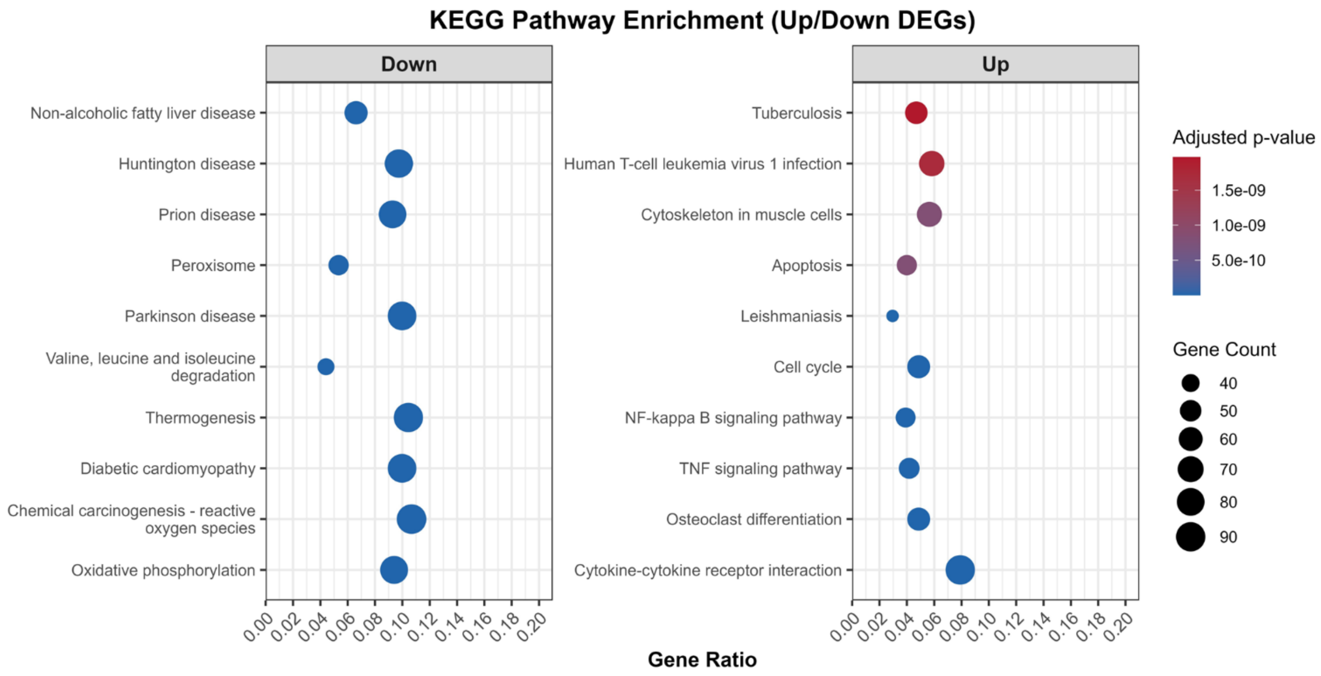


Supplementary Figure 5

(a)

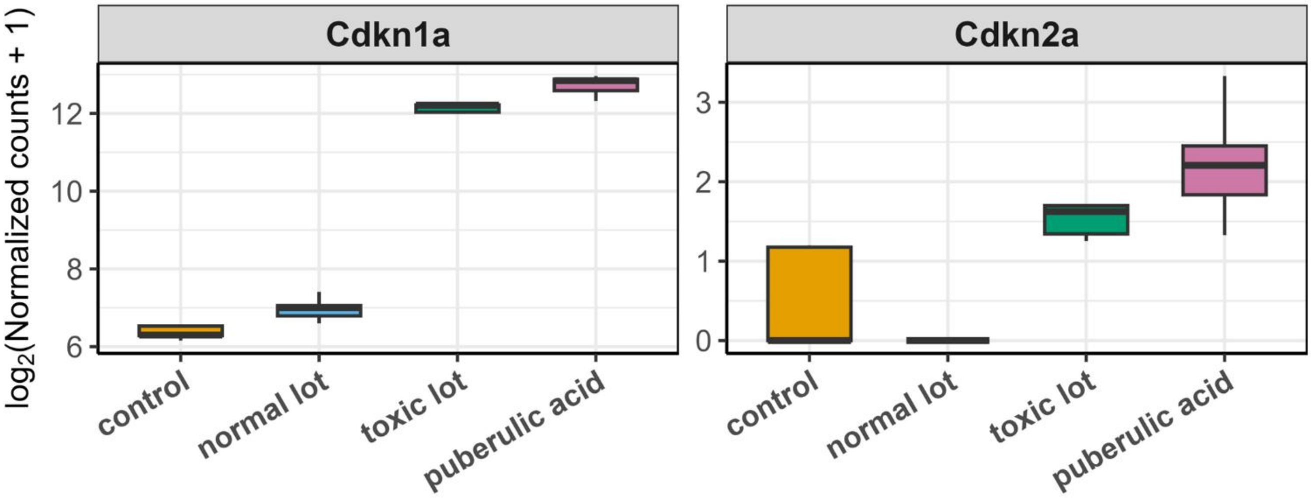


(b)

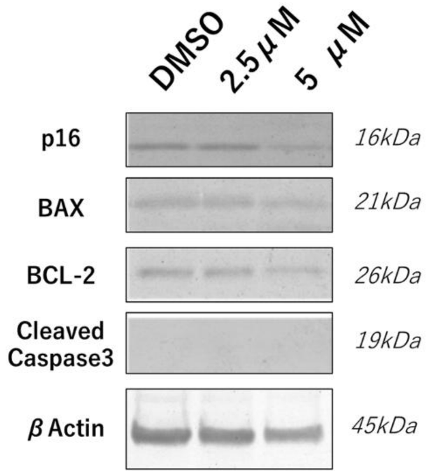


Supplementary Figure 6

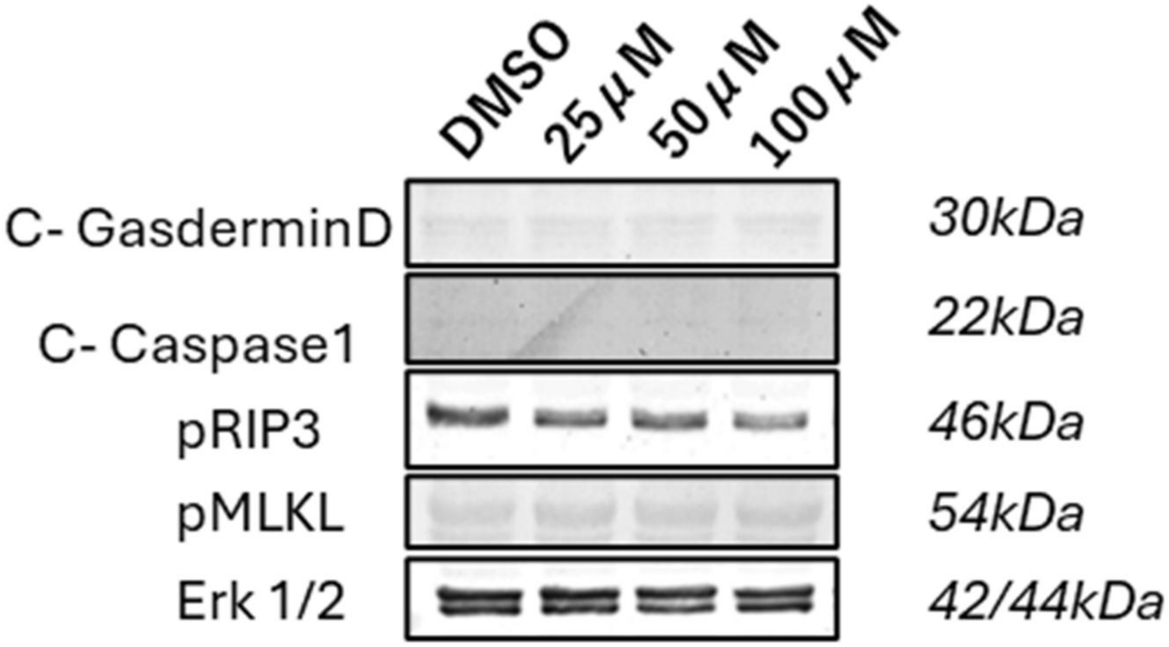


Supplementary Figure 7

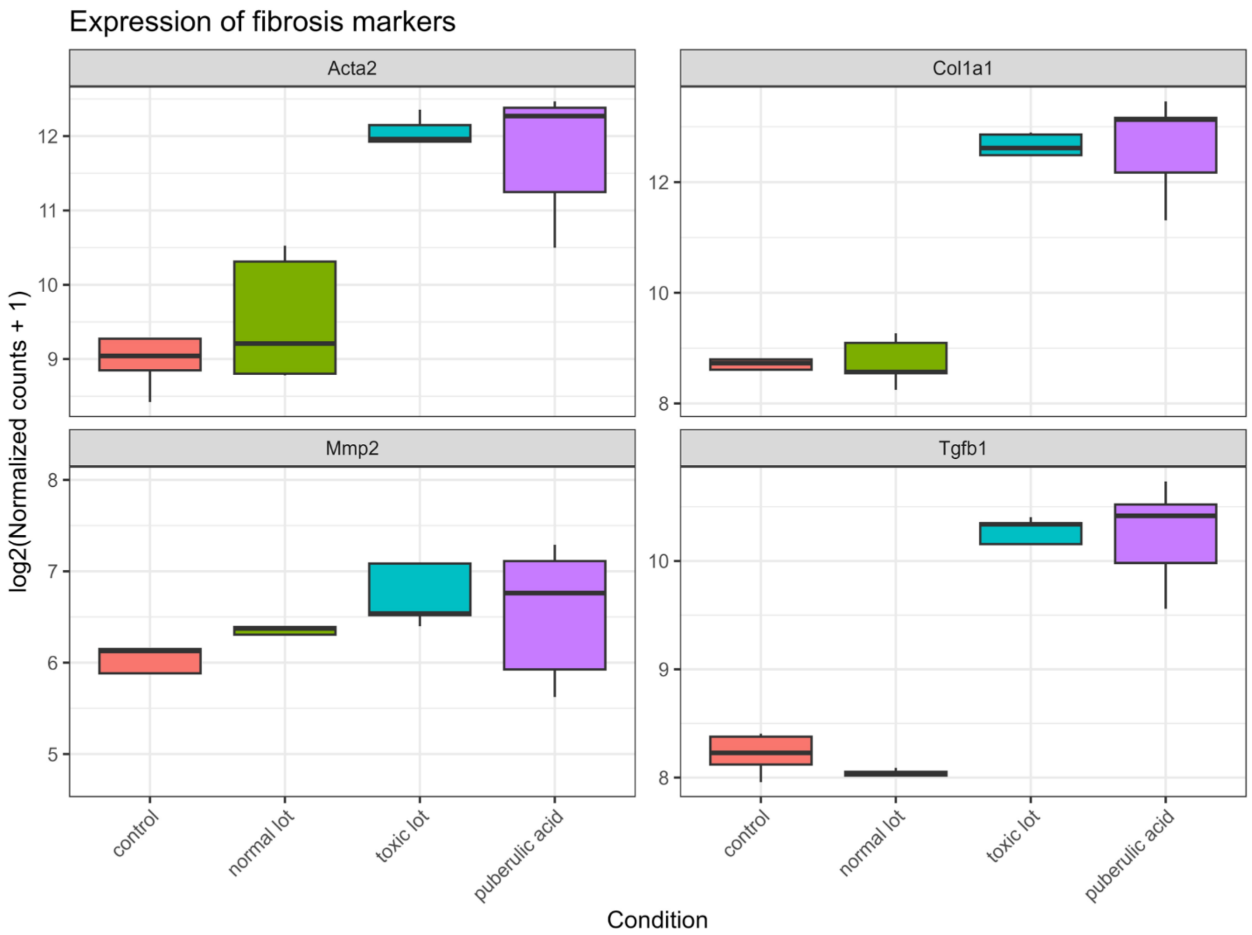


Supplementary Figure 8

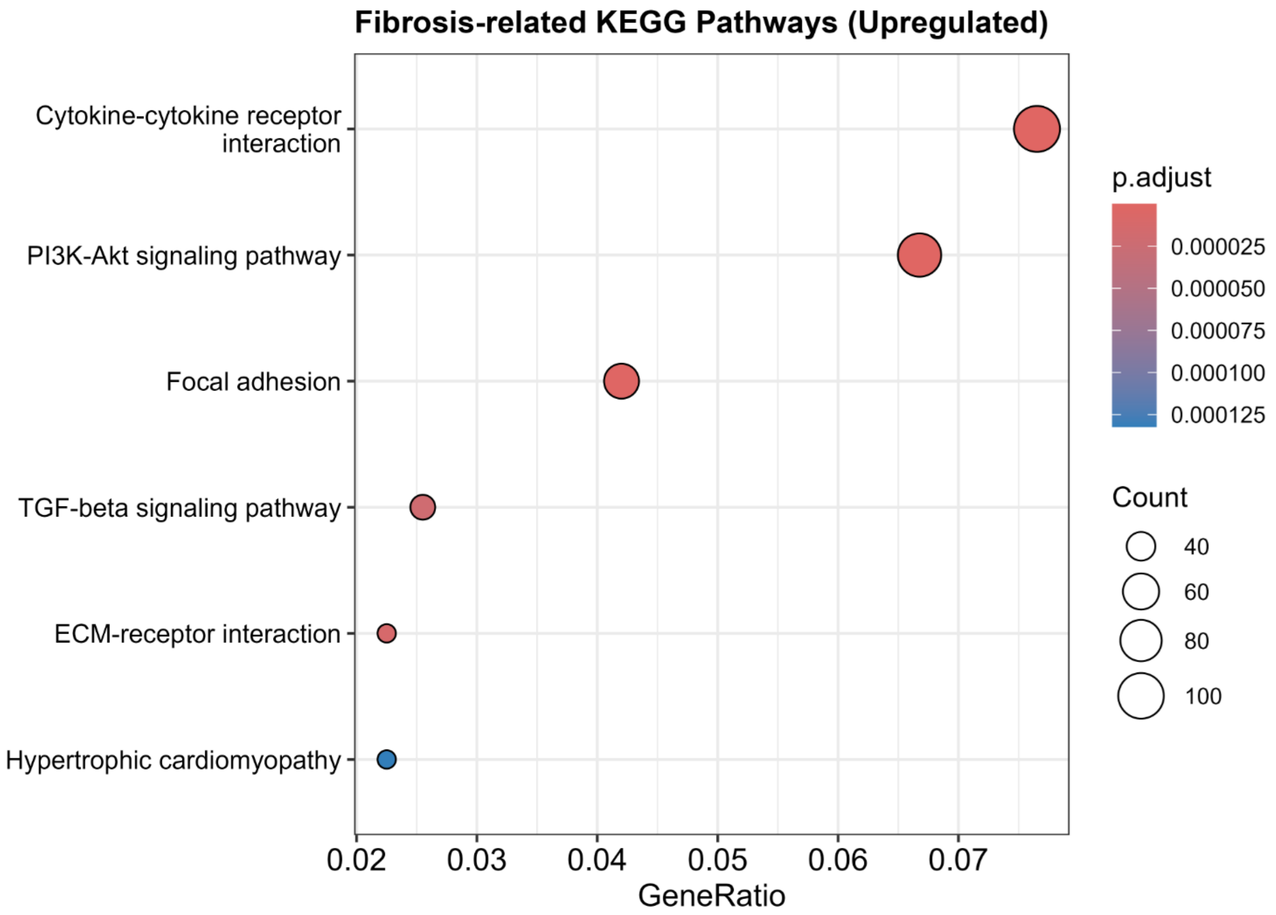


Supplementary Figure 9

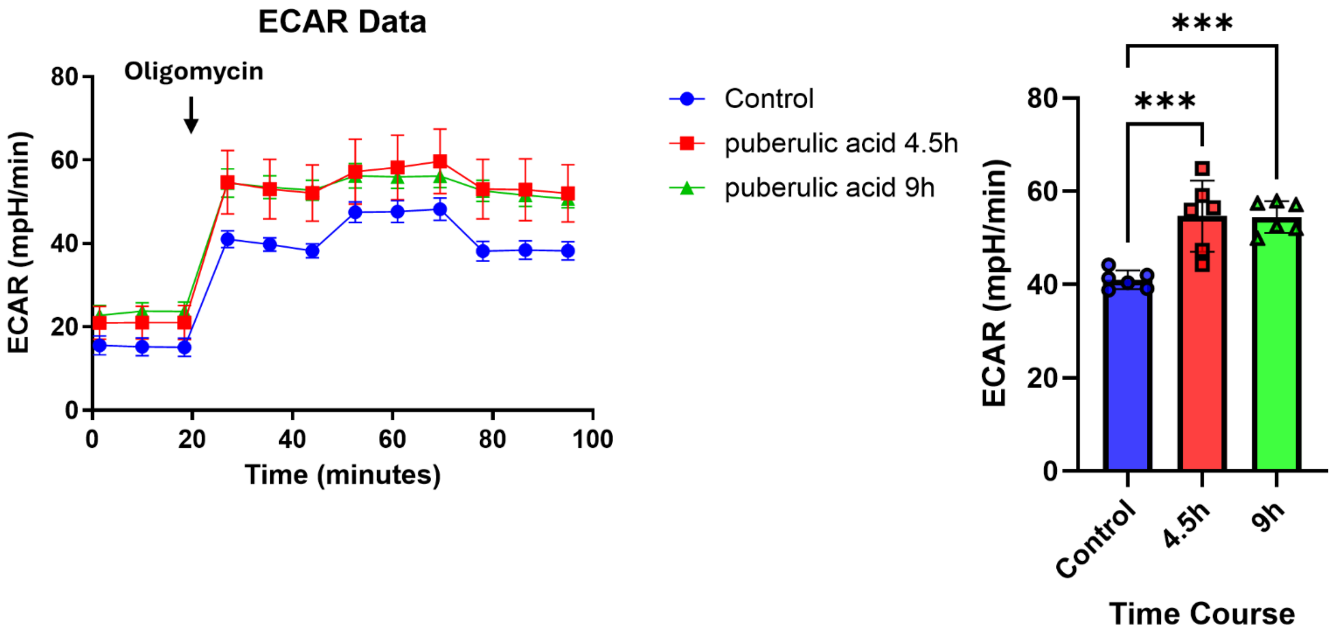


Supplementary Figure 10

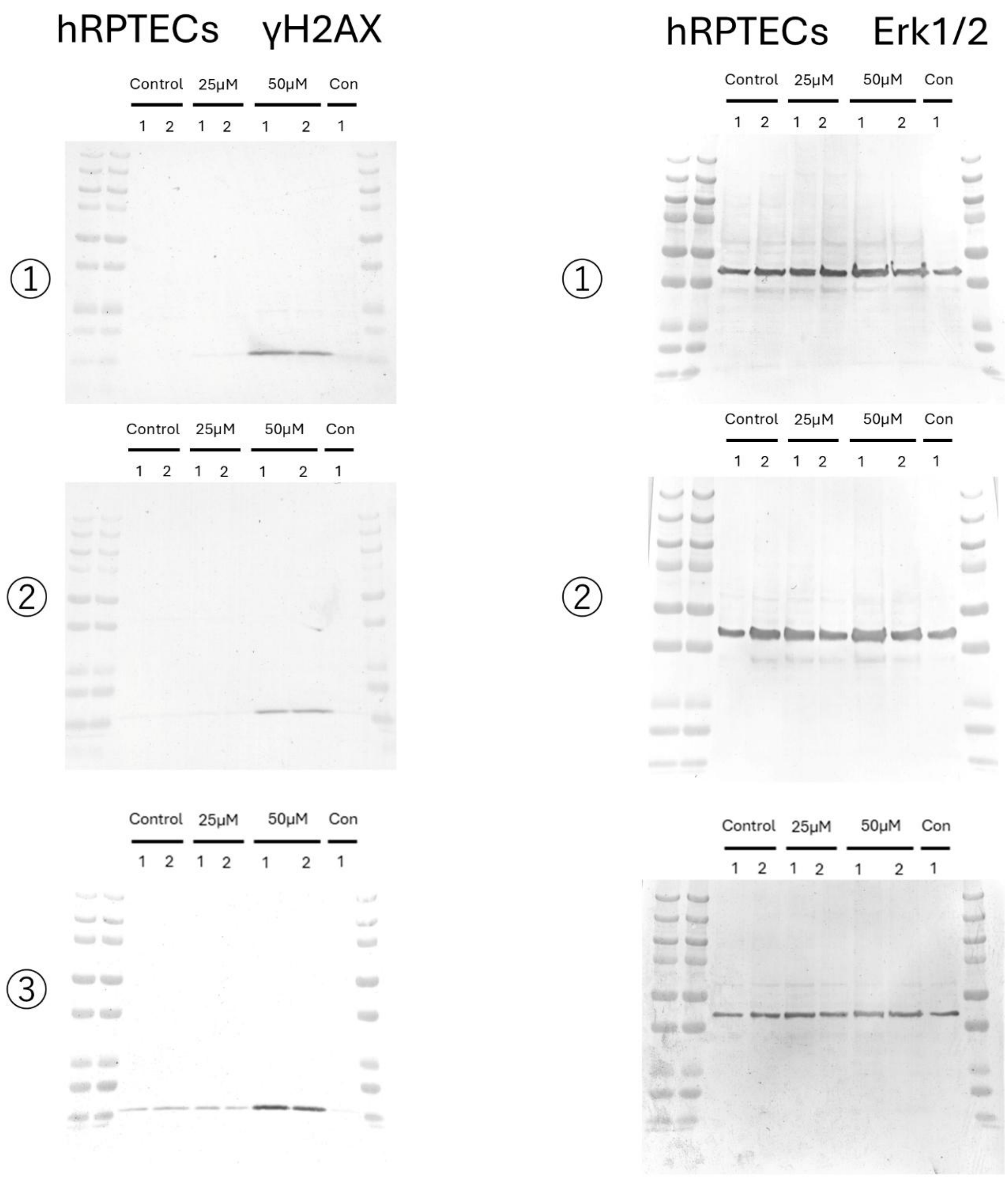


Full Blot of Figure 4b

tubuloids γH2AX

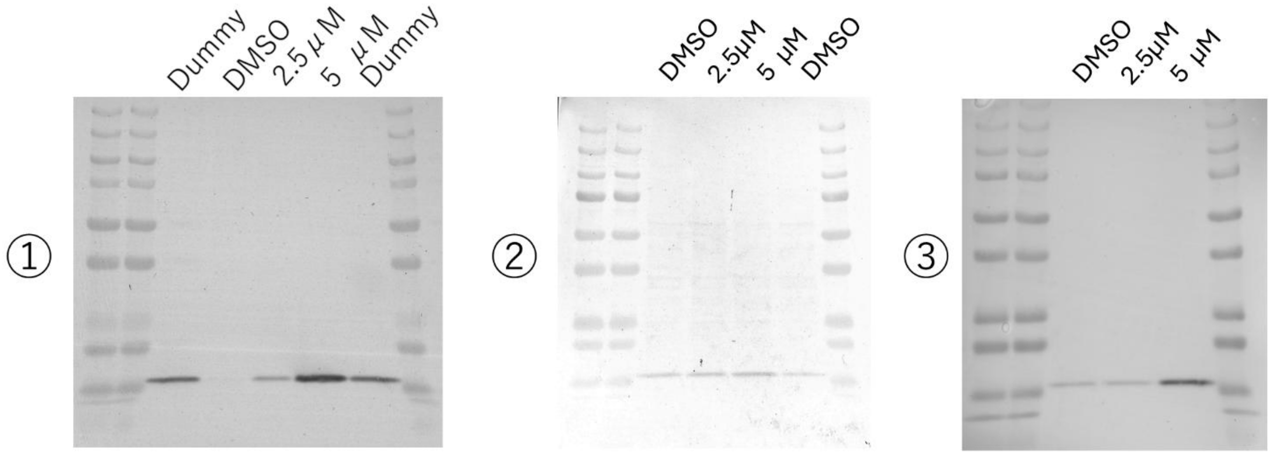


tubuloids beta Actin

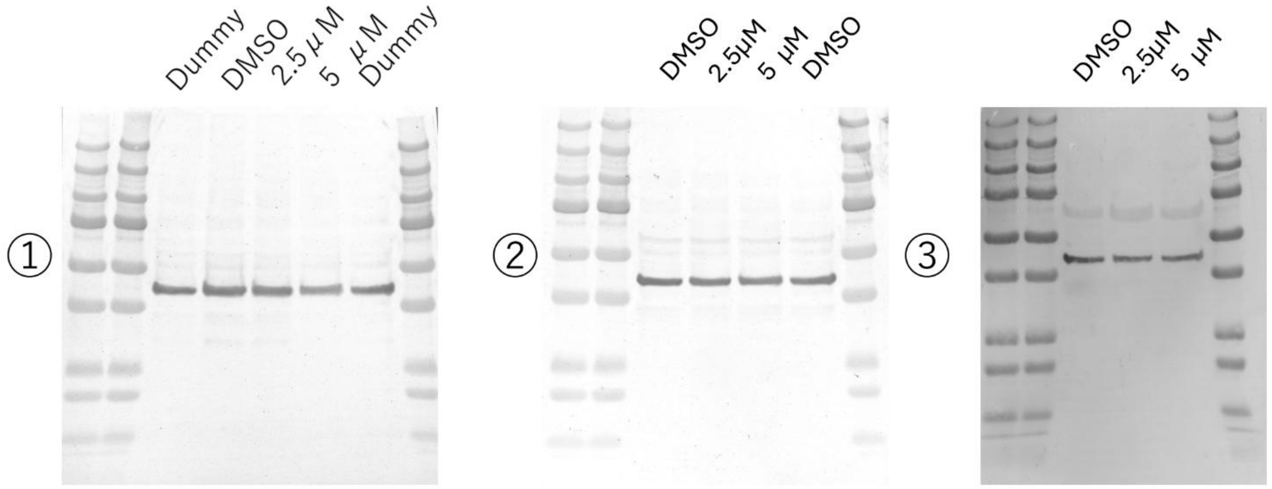


Full Blot of Figure 4c
